# Conversational Artificial Intelligence-Enabled Precision Oncology Reveals Context-Specific TGFβ and JAK/STAT Alterations in Pancreatic Cancer

**DOI:** 10.64898/2026.06.10.26355398

**Authors:** Fernando C. Diaz, Brigette Waldrup, Francisco G. Carranza, Sophia Manjarrez, Enrique Velazquez-Villarreal

**Affiliations:** Lineberger Comprehensive Cancer Center, University of North Carolina, Chapel Hill, NC, United States; City of Hope, Beckman Research Institute, Department of Integrative Translational Sciences, Duarte, CA; City of Hope Comprehensive Cancer Center, Duarte, CA

**Keywords:** Pancreatic Ductal Adenocarcinoma, Precision Oncology, Gemcitabine, Artificial Intelligence, LLM, Conversational AI, AI-Agents, TGFβ pathway, JAK/STAT pathway

## Abstract

**Background:** Pancreatic ductal adenocarcinoma (PDAC) is characterized by extensive molecular complexity, profound stromal remodeling, and limited responsiveness to systemic therapies. Although gemcitabine-based regimens remain widely utilized, the molecular pathways that influence treatment-associated biological variation are incompletely understood. The TGFβ and JAK/STAT signaling networks are recognized regulators of tumor progression, immune modulation, and therapeutic resistance; however, their genomic architecture in clinically stratified PDAC populations remains poorly defined.

**Methods:** We employed a conversational artificial intelligence–driven analytical framework to investigate TGFβ and JAK/STAT pathway alterations in a cohort of 184 PDAC patients. Clinical and molecular data were integrated to generate age- and treatment-stratified cohorts, enabling pathway-level and gene-level analyses according to gemcitabine exposure. Findings generated through AI-assisted interrogation were subsequently evaluated using conventional statistical approaches.

**Results:** TGFβ pathway alterations were identified in approximately one-quarter to one-third of tumors across clinical subgroups and demonstrated relatively stable frequencies regardless of age at diagnosis or gemcitabine treatment status. Gene-level analyses revealed that pathway disruption was predominantly driven by recurrent alterations in SMAD4, with additional low-frequency events involving TGFBR1 and TGFBR2. Notably, TGFBR2 mutations were significantly more frequent among late-onset PDAC patients receiving gemcitabine compared with untreated late-onset patients (8.8% vs. 1.4%; p = 0.04), suggesting a potential treatment-associated enrichment. In contrast, JAK/STAT pathway alterations were rare throughout the cohort, with only isolated mutations observed in pathway components including JAK1, JAK2, JAK3, STAT1, STAT3, and related regulatory genes. No significant differences in JAK/STAT alteration frequencies were identified according to age or treatment exposure.

**Conclusions:** TGFβ and JAK/STAT pathways exhibit distinct genomic architectures in PDAC. TGFβ pathway disruption represents a recurrent feature of disease biology, largely driven by SMAD4 alterations, while TGFBR2 enrichment in gemcitabine-treated late-onset tumors suggests a potential context-specific association worthy of further investigation. Conversely, genomic alterations within the JAK/STAT pathway are uncommon, indicating that pathway activity may be regulated predominantly through non-genomic mechanisms. These findings demonstrate the utility of conversational artificial intelligence agents for rapid, scalable, and clinically contextualized pathway interrogation and support future studies integrating multi-omic data to refine precision medicine strategies in PDAC.

## 1. Introduction

Pancreatic ductal adenocarcinoma (PDAC) remains one of the most lethal solid malignancies worldwide, with a 5-year survival rate that remains in the single digits despite advances in systemic therapy and molecular oncology (1–3). The aggressive clinical behavior of PDAC is driven by a combination of extensive molecular heterogeneity, profound stromal desmoplasia, immune evasion, and intrinsic resistance to conventional therapies. Although gemcitabine-based regimens continue to serve as a cornerstone of treatment for patients with advanced disease, therapeutic responses remain highly variable, and most patients eventually develop treatment resistance (1,4). These observations highlight the need to better understand the molecular pathways that influence treatment response and disease progression in clinically relevant patient subgroups.

Among the signaling networks implicated in PDAC biology, the transforming growth factor-beta (TGFβ) pathway occupies a particularly complex and context-dependent role. During early tumorigenesis, TGFβ signaling can exert tumor-suppressive effects by inhibiting epithelial cell proliferation. However, as tumors progress, the pathway frequently shifts toward a tumor-promoting function characterized by enhanced epithelial-to-mesenchymal transition (EMT), extracellular matrix remodeling, fibrosis, immune suppression, and metastatic dissemination (5–7). Alterations affecting key TGFβ pathway components, particularly SMAD4 and TGFβ receptors, occur in a substantial proportion of PDAC tumors and have been linked to aggressive disease behavior and therapeutic resistance (1,5,7).

Increasing evidence suggests that TGFβ signaling is also intimately involved in shaping the tumor microenvironment and modulating responses to gemcitabine. TGFβ-mediated activation of cancer-associated fibroblasts (CAFs) promotes extracellular matrix deposition, stromal remodeling, and reduced drug penetration, ultimately contributing to chemoresistance (8–12). Furthermore, TGFβ signaling regulates multiple stromal and immune cell populations that influence tumor progression, including fibroblasts, macrophages, and regulatory T cells (6,11,13,14). Experimental studies have demonstrated that inhibition of TGFβ signaling can enhance gemcitabine sensitivity, reduce metastatic potential, and improve therapeutic efficacy in preclinical PDAC models (10,15,16).

Parallel to TGFβ signaling, the Janus kinase/signal transducer and activator of transcription (JAK/STAT) pathway has emerged as a critical regulator of inflammation, immune evasion, tumor growth, and treatment resistance in PDAC (17–20). Activation of IL-6/JAK/STAT3 signaling promotes tumor cell proliferation, survival, cancer stemness, and remodeling of the tumor microenvironment, while also contributing to resistance against cytotoxic therapies (18,20–22). Moreover, gemcitabine itself has been shown to induce JAK/STAT-dependent upregulation of PD-L1 expression, potentially facilitating adaptive immune escape mechanisms during treatment (19,23,24). Several preclinical and clinical studies have therefore explored JAK inhibition as a strategy to enhance chemotherapy efficacy, although results have been variable and likely depend on the specific biological context and patient population examined (17,22,25).

Importantly, growing evidence indicates that substantial crosstalk exists between TGFβ and JAK/STAT signaling networks. TGFβ-mediated signaling can induce STAT3 activation through both canonical and non-canonical mechanisms, while inflammatory cytokines regulated by JAK/STAT signaling can reinforce TGFβ-driven fibrosis, immune suppression, and tumor progression (5,6,18,26). These interactions create a dynamic signaling landscape that influences therapeutic response, stromal remodeling, and disease evolution. Understanding the genomic architecture of these pathways may therefore provide valuable insights into biologically distinct PDAC subgroups and reveal opportunities for precision medicine-based intervention.

Despite the growing recognition of pathway-driven tumor heterogeneity, comprehensive interrogation of clinically annotated genomic datasets remains challenging. Traditional bioinformatics workflows often require extensive programming expertise and lack the flexibility to rapidly generate, refine, and test clinically relevant hypotheses across multiple patient subgroups. These limitations are particularly relevant when evaluating age-dependent and treatment-dependent molecular patterns that may influence therapeutic outcomes.

To address this challenge, we applied a conversational artificial intelligence framework to investigate TGFβ and JAK/STAT pathway alterations in PDAC. Building upon our previously reported AI-HOPE precision oncology platform and pathway-specific conversational agents (27) this approach enables dynamic cohort construction, rapid genomic interrogation, and real-time hypothesis testing using natural language interactions. Using age- and treatment-stratified PDAC cohorts, we evaluated the distribution of TGFβ and JAK/STAT pathway alterations and explored their potential clinical significance in the context of gemcitabine exposure. Through this precision medicine-oriented strategy, we sought to identify context-specific molecular dependencies that may contribute to treatment response and inform future pathway-targeted therapeutic approaches in PDAC.

## 2. Results

### 2.1 Demographic and Clinical Characteristics of the Study Population

The study included 184 patients diagnosed with PDAC, all of whom had molecular data generated from primary tumor specimens. To evaluate the clinical and biological relevance of pathway alterations according to age at diagnosis, the cohort was stratified into early-onset PDAC (EO-PDAC; <50 years) and late-onset PDAC (LO-PDAC; ≥50 years) groups (Table 1).

**Table 1.** Overview of Baseline Demographic and Clinical Characteristics of Early-Onset and Late-Onset PDAC Patients.

| Clinical Feature | Early-Onset<br>n (%) | Late-Onset<br>n (%) |
| --- | --- | --- |
| **Treatment** |  |  |
| Treated with Gemcitabine | 15 (75.0%) | 91 (55.5%) |
| Not Treated with Gemcitabine | 5 (25.0%) | 73 (44.5%) |
| **Cancer Type** |  |  |
| Pancreatic Adenocarcinoma | 20 (100.0%) | 164 (100.0%) |
| **Sex** |  |  |
| Male | 11 (55.0%) | 90 (54.9%) |
| Female | 9 (45.0%) | 74 (45.1%) |
| **Sample Type** |  |  |
| Primary Tumor | 20 (100.0%) | 164 (100.0%) |
| **Stage at Diagnosis** |  |  |
| Stage I | 2 (10.0%) | 19 (11.6%) |
| Stage II | 17 (85.0%) | 134 (81.7%) |
| Stage III | 1 (5.0%) | 4 (2.4%) |
| Stage IV | 0 (0.0%) | 5 (3.0%) |
| NA | 0 (0.0%) | 2 (1.2%) |
| **Ethnicity** |  |  |
| Hispanic or Latino | 1 (5.0%) | 4 (2.4%) |
| Not Hispanic or Latino | 14 (70.0%) | 122 (74.4%) |
| Unknown | 5 (25.0%) | 38 (23.2%) |

Among the study population, 20 patients (10.9%) were classified as EO-PDAC, whereas 164 patients (89.1%) were diagnosed with LO-PDAC. Gemcitabine-based therapy was administered to the majority of patients in both age groups, including 15 EO-PDAC patients (75.0%) and 91 LO-PDAC patients (55.5%). Conversely, 5 EO-PDAC patients (25.0%) and 73 LO-PDAC patients (44.5%) did not receive gemcitabine treatment.

The cohort demonstrated a balanced sex distribution across age groups. In the EO-PDAC subgroup, 11 patients (55.0%) were male and 9 (45.0%) were female. Similarly, the LO-PDAC subgroup included 90 male patients (54.9%) and 74 female patients (45.1%). All molecular analyses were performed using primary pancreatic adenocarcinoma specimens, ensuring consistency in tissue source and minimizing variability associated with metastatic or recurrent disease samples.

Most tumors were diagnosed at stage II disease regardless of age at onset. Among EO-PDAC patients, 17 cases (85.0%) were classified as stage II, while 2 patients (10.0%) presented with stage I disease and 1 patient (5.0%) with stage III disease. No stage IV tumors were observed in the EO-PDAC group. In contrast, the LO-PDAC cohort included 19 stage I tumors (11.6%), 134 stage II tumors (81.7%), 4 stage III tumors (2.4%), and 5 stage IV tumors (3.0%). Staging information was unavailable for only two LO-PDAC patients (1.2%).

Ethnicity data indicated that most patients were classified as not Hispanic or Latino. This group represented 70.0% of EO-PDAC patients and 74.4% of LO-PDAC patients. Hispanic or Latino individuals accounted for 5.0% and 2.4% of EO-PDAC and LO-PDAC cases, respectively. Ethnicity information was unavailable for 25.0% of EO-PDAC patients and 23.2% of LO-PDAC patients.

Overall, the cohort reflects a predominantly late-onset PDAC population with substantial representation of gemcitabine-treated patients and a predominance of stage II disease at diagnosis. The comparable distributions of sex and clinical stage between EO-PDAC and LO-PDAC groups provide an appropriate framework for investigating age-dependent molecular alterations and their potential associations with treatment response and clinical outcomes.

### 2.2 Context-Dependent Distribution of TGFβ and JAK/STAT Pathway Alterations Across Age and Gemcitabine Treatment Groups

Stratified analyses according to age at diagnosis and gemcitabine treatment exposure (Table 2a–d) revealed distinct patterns of pathway disruption between the TGFβ and JAK/STAT signaling networks. While TGFβ pathway alterations were relatively common across multiple clinical contexts, JAK/STAT pathway alterations were rare throughout the cohort, suggesting fundamentally different roles for these pathways in PDAC biology.

**Table 2.**
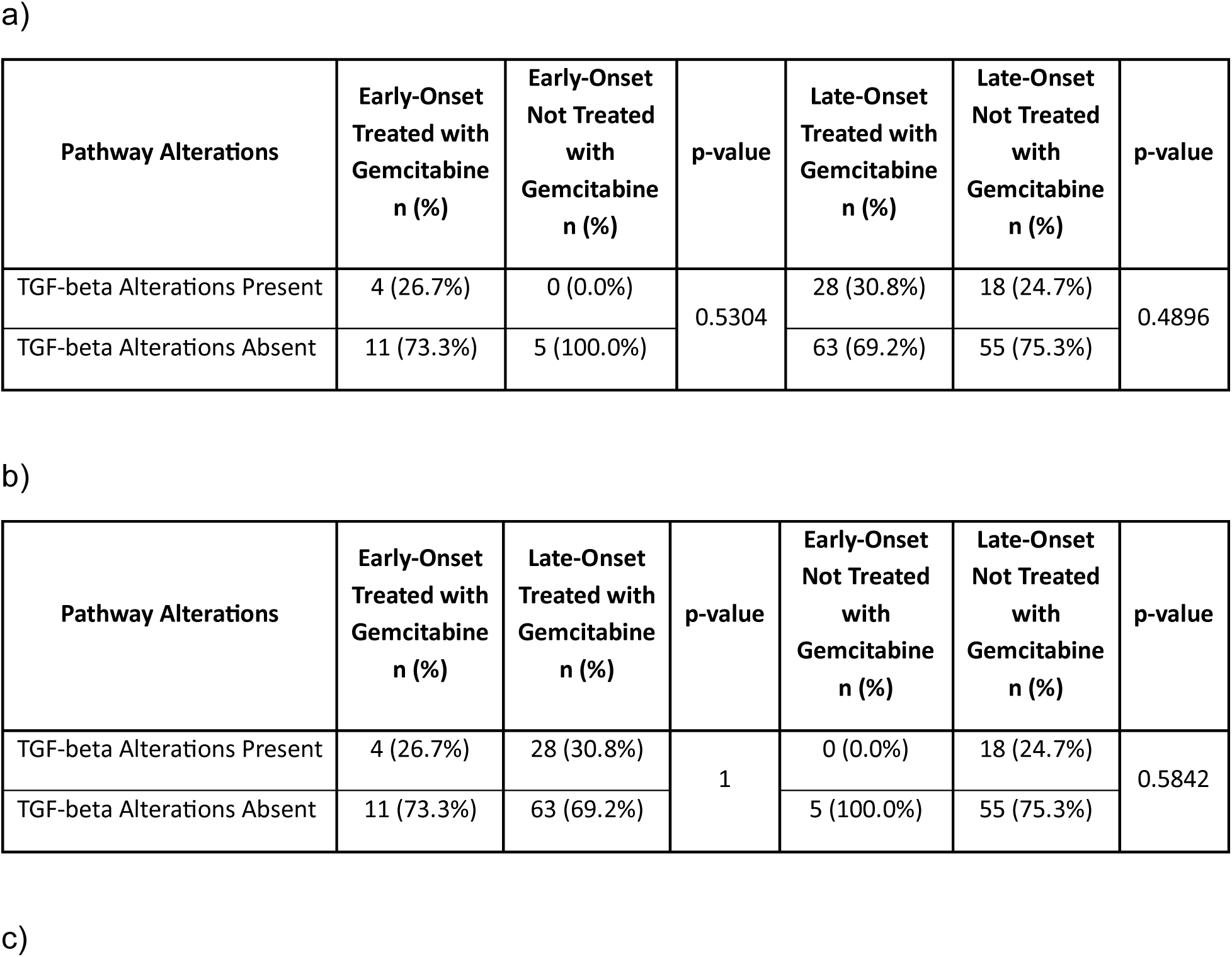
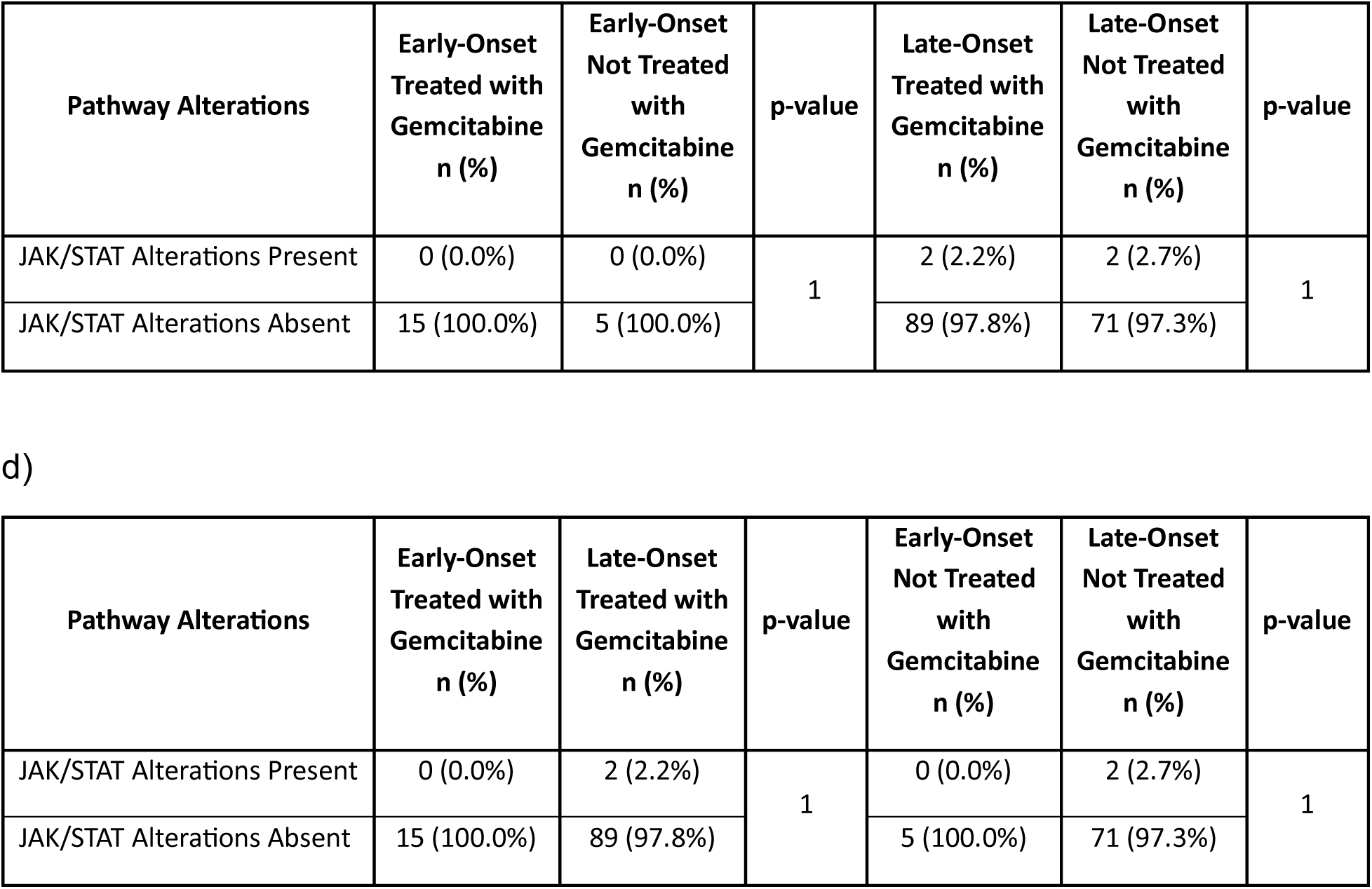
Distribution of TGFβ and JAK/STAT Pathway Alterations Across Age- and Treatment-Stratified PDAC Subgroups. This table summarizes the frequency of TGFβ and JAK/STAT pathway alterations across PDAC cohorts stratified by age at diagnosis (early-onset vs. late-onset) and gemcitabine treatment exposure. The analysis is organized into four complementary panels to facilitate pathway-specific and clinically contextual interpretation: (a) TGFβ pathway alterations comparing gemcitabine-treated and non-gemcitabine-treated tumors within early-onset and late-onset PDAC cohorts; (b) TGFβ pathway alterations comparing early-onset and late-onset disease within each treatment stratum; (c) JAK/STAT pathway alterations comparing gemcitabine-treated and non-treated tumors within early-onset and late-onset PDAC cohorts; and (d) JAK/STAT pathway alterations comparing early-onset and late-onset PDAC within gemcitabine-treated and non-treated groups. P-values were calculated to assess differences in alteration frequencies between the respective comparison groups.

#### 2.2.1 TGFβ pathway alterations across age and treatment strata

TGFβ pathway alterations were observed in both early-onset and late-onset PDAC and demonstrated moderate prevalence across treatment groups. Within early-onset PDAC, TGFβ alterations were identified in 26.7% of gemcitabine-treated tumors, whereas no alterations were detected among non-treated patients (0.0%). However, this difference did not reach statistical significance (p = 0.53), likely reflecting the limited sample size of the early-onset subgroup (Table 2a).

Among late-onset PDAC patients, TGFβ pathway alterations were detected in 30.8% of gemcitabine-treated tumors and 24.7% of non-treated tumors. Similar to the early-onset cohort, no statistically significant difference was observed between treatment groups (p = 0.48). These findings suggest that TGFβ pathway dysregulation occurs frequently in PDAC regardless of gemcitabine exposure and may represent an intrinsic biological feature of the disease rather than a treatment-associated event.

Comparisons across age groups within treatment strata further supported the consistency of TGFβ pathway alteration frequencies. Among gemcitabine-treated patients, alteration rates were comparable between early-onset (26.7%) and late-onset (30.8%) PDAC (p = 1.0). Likewise, among patients not treated with gemcitabine, no significant differences were observed between age groups, although alterations were detected only in late-onset tumors (24.7%) and not in early-onset cases (p = 0.58) (Table 2b).

Overall, TGFβ pathway alterations emerged as a recurrent molecular feature across age and treatment contexts, with approximately one-quarter to one-third of tumors exhibiting pathway disruption. The stability of these frequencies across clinical subgroups suggests that TGFβ signaling may represent a broadly relevant therapeutic and biological pathway in PDAC.

#### 2.2.2 JAK/STAT pathway alterations across age and treatment strata

In contrast to TGFβ signaling, JAK/STAT pathway alterations were uncommon throughout the study cohort. No JAK/STAT alterations were identified in early-onset PDAC regardless of gemcitabine treatment status, with all tumors classified as pathway unaltered (Table 2c).

Within late-onset PDAC, JAK/STAT pathway alterations were detected in only 2.2% of gemcitabine-treated tumors and 2.7% of non-treated tumors. No statistically significant differences were observed between treatment groups (p = 1.0). Similarly, age-stratified comparisons demonstrated no significant variation in pathway alteration frequencies among either gemcitabine-treated or non-treated patients (Table 2d).

The uniformly low prevalence of JAK/STAT pathway alterations across all clinical contexts suggests that direct genomic disruption of this signaling network is infrequent in PDAC. Nevertheless, given the established role of cytokine-mediated JAK/STAT signaling in tumor progression and immune regulation, these findings do not exclude the possibility of pathway activation through transcriptional, epigenetic, or microenvironmental mechanisms that are not captured by genomic alteration analyses alone.

These results highlight a clear contrast between the two pathways evaluated. TGFβ pathway alterations were relatively common and consistently observed across age and treatment groups, whereas JAK/STAT pathway alterations were rare irrespective of clinical context. These observations provide a foundation for subsequent survival and precision medicine analyses performed using conversational artificial intelligence agents designed to interrogate clinically relevant molecular subgroups in PDAC.

### 2.3 Gene-Level Landscape of TGFβ and JAK/STAT Signaling Pathways

Given the distinct pathway-level patterns observed for TGFβ and JAK/STAT signaling (Section 2.2), we next examined individual pathway components to identify gene-specific alterations that may contribute to PDAC heterogeneity and precision medicine stratification (Tables S1–S8). Gene-level analyses revealed fundamentally different architectures between the two pathways. The TGFβ pathway was characterized by recurrent alterations concentrated within a limited number of canonical signaling genes, particularly SMAD4, whereas the JAK/STAT pathway demonstrated an exceptionally low mutational burden with sparse alterations distributed across multiple pathway members. These findings indicate that pathway-level observations are largely driven by distinct underlying molecular mechanisms and highlight the value of conversational AI-assisted interrogation of genomic datasets for identifying clinically relevant patterns.

#### 2.3.1 TGFβ gene-level landscape is dominated by SMAD4 alterations

Within the TGFβ signaling network, SMAD4 emerged as the predominant altered gene across all clinical subgroups. In early-onset PDAC, SMAD4 alterations were detected in 20.0% of gemcitabine-treated tumors but were absent in patients who did not receive gemcitabine, although this difference did not reach statistical significance (p=0.53) (Table S1). A small number of additional alterations were observed in TGFBR2 (6.7%) among gemcitabine-treated early-onset tumors, while all remaining TGFβ pathway genes were unaltered. These findings suggest that TGFβ pathway disruption in early-onset disease is primarily driven by alterations affecting core signal transduction components rather than widespread pathway dysregulation.

In late-onset PDAC, TGFBR2 mutations were significantly more frequent in late-onset PDAC patients treated with gemcitabine compared to those not treated (8.8% vs. 1.4%, p = 0.04), suggesting a potential association between TGFβ signaling disruption and therapeutic exposure in this subgroup. SMAD4 remained the most frequently altered gene, occurring in 22.0% of gemcitabine-treated tumors and 19.2% of non-treated tumors. Additional low-frequency alterations involved TGFBR2 (8.8% and 1.4%, respectively), TGFBR1 (4.4% in treated tumors), and isolated mutations affecting BMP family members and receptor-associated genes. No individual gene demonstrated significant differences according to treatment status, indicating that TGFβ pathway disruption is largely independent of gemcitabine exposure (Tables S2–S4).

Comparisons between age groups further demonstrated the remarkable consistency of TGFβ pathway architecture. Among gemcitabine-treated patients, SMAD4 alterations occurred in 20.0% of early-onset tumors and 22.0% of late-onset tumors (p=1.0), while TGFBR2 alterations were identified in 6.7% and 8.8% of cases, respectively (Table S3). Similar findings were observed in the non-treated cohort, where SMAD4 alterations were present only in late-onset tumors but did not differ significantly between age groups (p=0.5786) (Table S4). Collectively, these data indicate that SMAD4-driven disruption represents a conserved feature of PDAC regardless of age at diagnosis or treatment exposure.

#### 2.3.2 JAK/STAT gene-level landscape demonstrates extremely low mutational burden

In contrast to the TGFβ pathway, genomic alterations affecting JAK/STAT signaling were exceedingly uncommon throughout the cohort. No JAK/STAT pathway mutations were detected in early-onset PDAC, regardless of gemcitabine treatment status (Table S5). All evaluated pathway members, including JAK1, JAK2, JAK3, STAT1, STAT3, STAT5A, STAT5B, STAT6, PIAS family genes, PTPRC, and SOCS1, were unaltered in both treated and untreated early-onset tumors. These findings are consistent with the absence of pathway-level JAK/STAT alterations observed in early-onset disease.

Late-onset PDAC demonstrated only sporadic alterations distributed across multiple JAK/STAT pathway components. Individual mutations in JAK1, JAK2, JAK3, PIAS1, PIAS2, PIAS3, PTPRC, STAT1, STAT3, STAT5A, STAT5B, and STAT6 were each detected in approximately 1.1% of tumors, whereas SOCS1 remained unaltered across all patients (Table S6). Notably, the frequency of these mutations was virtually identical between gemcitabine-treated and non-treated groups, with no statistically significant differences identified for any gene. This pattern suggests that JAK/STAT pathway alterations occur as isolated events rather than recurrent driver mutations.

Age-based comparisons reinforced the rarity of JAK/STAT pathway disruption. Whereas no mutations were identified in early-onset PDAC, late-onset tumors harbored only occasional single-gene alterations, each occurring in approximately 1% of cases and without statistically significant enrichment in any subgroup (Tables S7 and S8). The absence of recurrently altered genes indicates that direct genomic activation of the JAK/STAT pathway is uncommon in PDAC and may not represent the principal mechanism through which this signaling network contributes to disease biology.

#### 2.3.3 Integrated interpretation

Taken together, gene-level analyses reveal two distinct patterns of pathway dysregulation in PDAC. The TGFβ pathway is characterized by recurrent alterations concentrated in a small number of canonical signaling components, particularly SMAD4, with remarkably consistent frequencies across age groups and treatment contexts. This finding supports a stable and conserved role for TGFβ pathway disruption in PDAC pathogenesis. In contrast, the JAK/STAT pathway exhibits minimal genomic alteration, with only isolated low-frequency mutations scattered across multiple genes and no evidence of recurrent driver events.

These results suggest that TGFβ signaling alterations may be directly linked to tumor-intrinsic genomic mechanisms, whereas JAK/STAT pathway activity in PDAC is likely influenced by non-genomic processes such as cytokine signaling, transcriptional regulation, or tumor microenvironment interactions. From a precision medicine perspective, the predominance of SMAD4 alterations highlights the TGFβ axis as a potentially informative biomarker and therapeutic target, whereas genomic assessment of JAK/STAT pathway genes alone may provide limited discriminatory value for patient stratification. The ability of conversational artificial intelligence agents to rapidly identify and contextualize these gene-level patterns further demonstrates their utility for precision oncology research and hypothesis generation.

### 2.4 Context-Specific Survival Impact of TGF-b Pathway Alterations

Kaplan–Meier analyses were performed to investigate the association between TGFβ pathway dysregulation and overall survival in PDAC patients categorized by age at diagnosis and receipt of gemcitabine therapy (Figure 1a–d).

**Figure 1.**
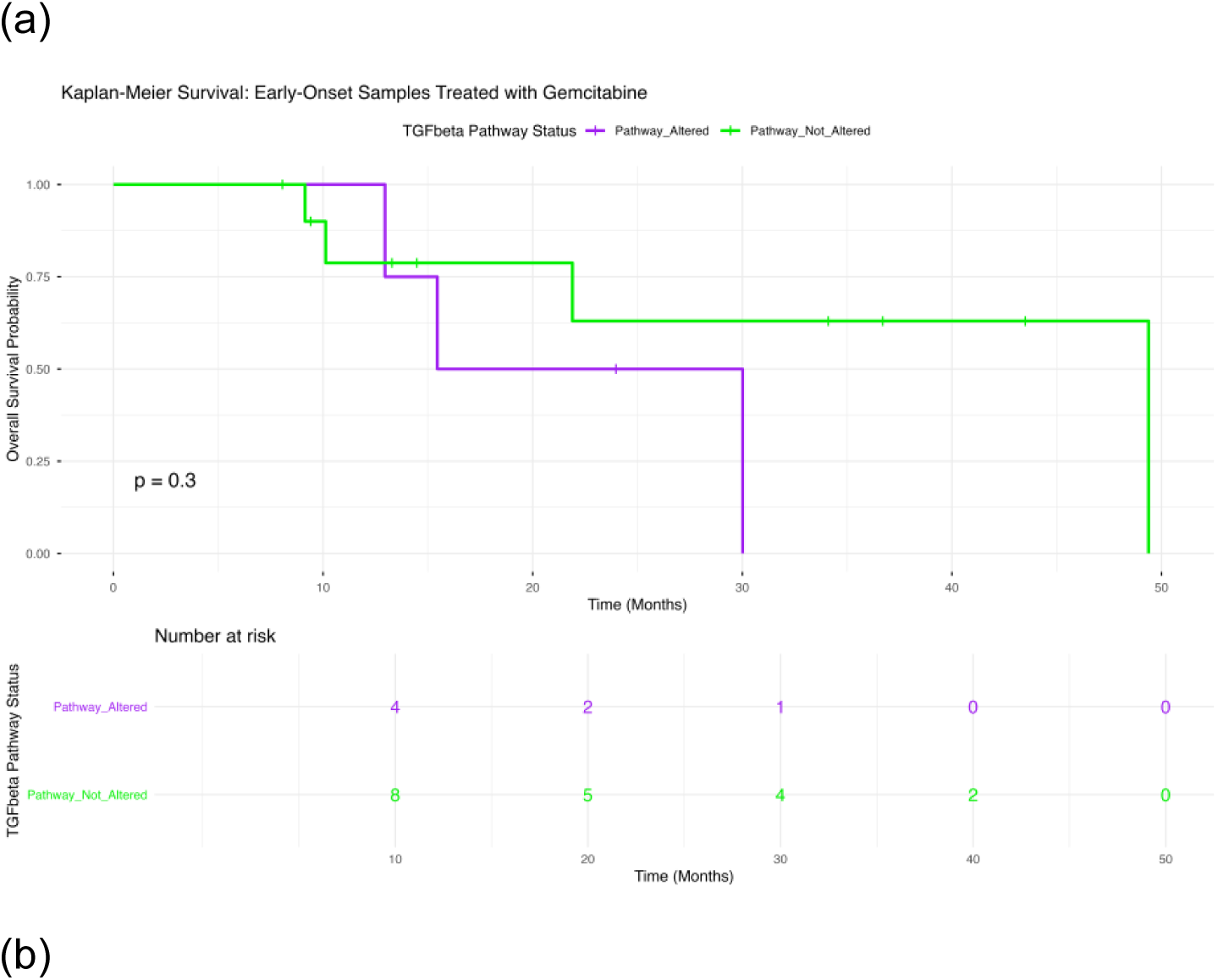
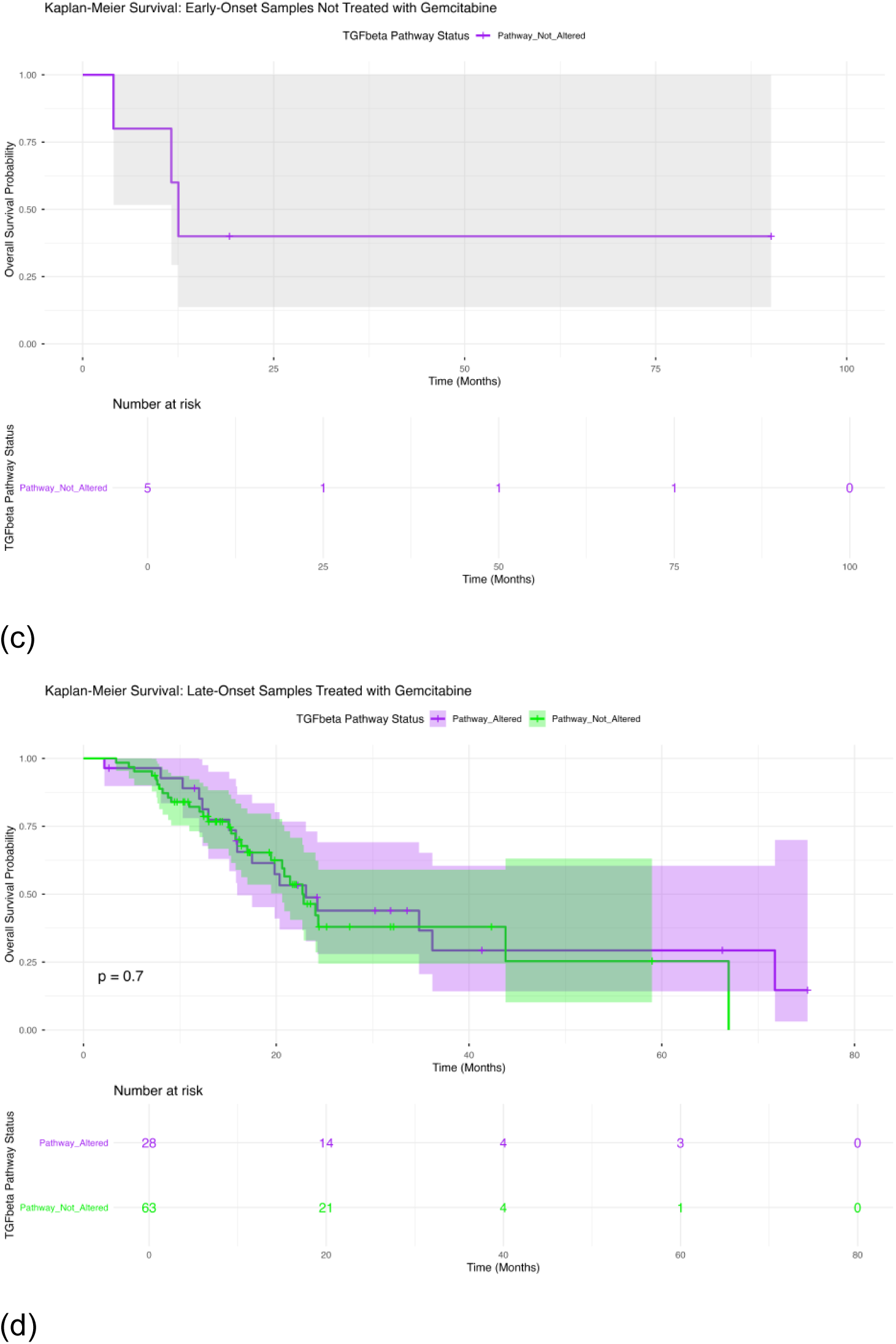
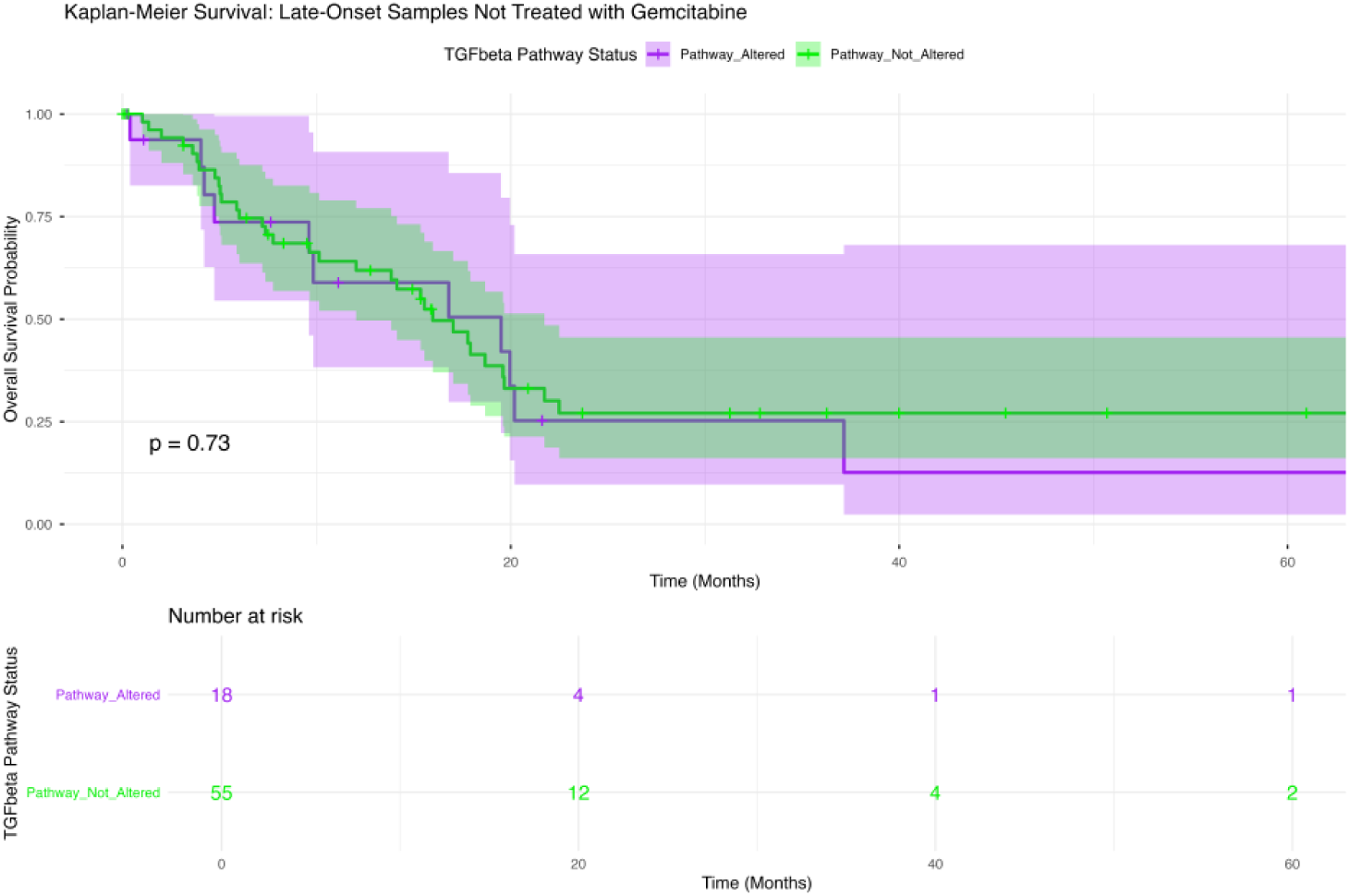
Kaplan–Meier survival analysis according to TGFβ and JAK/STAT pathway alteration status across age- and treatment-specific PDAC subgroups. Overall survival was evaluated in PDAC patients stratified by molecular alterations in the TGFβ and JAK/STAT signaling pathways. Survival curves compare patients harboring pathway alterations with those lacking detectable alterations within four clinically defined subgroups: (a) early-onset PDAC (<50 years) treated with gemcitabine, (b) early-onset PDAC (<50 years) not treated with gemcitabine, (c) late-onset PDAC (≥50 years) treated with gemcitabine, and (d) late-onset PDAC (≥50 years) not treated with gemcitabine. Shaded regions indicate 95% confidence intervals. Numbers at risk are displayed beneath each plot. These analyses were generated using the AI-HOPE conversational artificial intelligence framework to investigate the prognostic relevance of pathway-specific molecular alterations in precision medicine–oriented PDAC subgroups.

#### 2.4.1 Early-Onset PDAC Under Gemcitabine Exposure

Among patients with early-onset PDAC who received gemcitabine-based therapy (Figure 1a), no statistically significant association was observed between TGFβ pathway alteration status and overall survival (p = 0.3). Patients harboring TGFβ pathway alterations exhibited a decline in survival beginning approximately 12–15 months after diagnosis, whereas those without detectable alterations demonstrated a more gradual decrease in survival probability over time. Despite these differences in survival trajectories, the curves remained largely overlapping throughout follow-up, indicating limited prognostic discrimination based on TGFβ pathway status in this subgroup. Interpretation of these findings should be made with caution given the small number of patients at risk and the limited sample size, which may reduce statistical power to detect clinically meaningful survival differences.

#### 2.4.2 Early-Onset PDAC Without Gemcitabine Exposure

Among early-onset PDAC patients who did not receive gemcitabine therapy (Figure 1b), evaluation of the association between TGFβ pathway alteration status and overall survival was limited by the absence of patients harboring detectable TGFβ pathway alterations within this subgroup. Consequently, survival estimates were derived exclusively from patients without pathway alterations, precluding a direct comparison between molecular groups. The observed survival curve demonstrated an initial decline during the first year of follow-up, followed by a prolonged plateau with approximately 40% survival probability maintained over time. Given the very small cohort size and lack of a comparison group, these findings should be interpreted with caution and do not allow definitive conclusions regarding the prognostic significance of TGFβ pathway alterations in untreated early-onset PDAC.

#### 2.4.3 Late-Onset PDAC Under Gemcitabine Exposure

Among patients with late-onset PDAC treated with gemcitabine (Figure 1c), TGFβ pathway alteration status was not significantly associated with overall survival (p = 0.7). Survival trajectories for tumors with and without detectable TGFβ pathway alterations were largely comparable throughout the follow-up period, with both groups demonstrating a progressive decline in survival probability over time. Although minor fluctuations were observed at later time points, no consistent survival advantage or disadvantage was evident for either molecular subgroup. The substantial overlap of the confidence intervals further suggests that TGFβ pathway status does not meaningfully influence clinical outcomes in gemcitabine-treated late-onset PDAC within this cohort. These findings indicate that TGFβ pathway alterations alone may have limited prognostic value in this patient population.

#### 2.4.4 Late-Onset PDAC Without Gemcitabine Exposure

In late-onset PDAC patients who did not receive gemcitabine treatment (Figure 1d), TGFβ pathway alteration status was not significantly associated with overall survival (p = 0.73). Survival curves for patients with and without TGFβ pathway alterations followed similar trajectories throughout the observation period, exhibiting substantial overlap across most time points. Although patients without detectable TGFβ alterations demonstrated a modest survival advantage during intermediate follow-up, this difference was not sustained and did not translate into a statistically significant separation between groups. The wide and overlapping confidence intervals at later time points further underscore the uncertainty of these estimates. Collectively, these findings suggest that TGFβ pathway alterations do not exert a measurable prognostic effect in late-onset PDAC patients who were not exposed to gemcitabine therapy.

### 2.5 Survival Outcomes Associated with JAK/STAT Pathway Dysregulation

To assess the clinical relevance of JAK/STAT pathway alterations, overall survival was examined across age- and treatment-defined PDAC subgroups using Kaplan–Meier methodology (Figure 2a–d).

**Figure 2.**
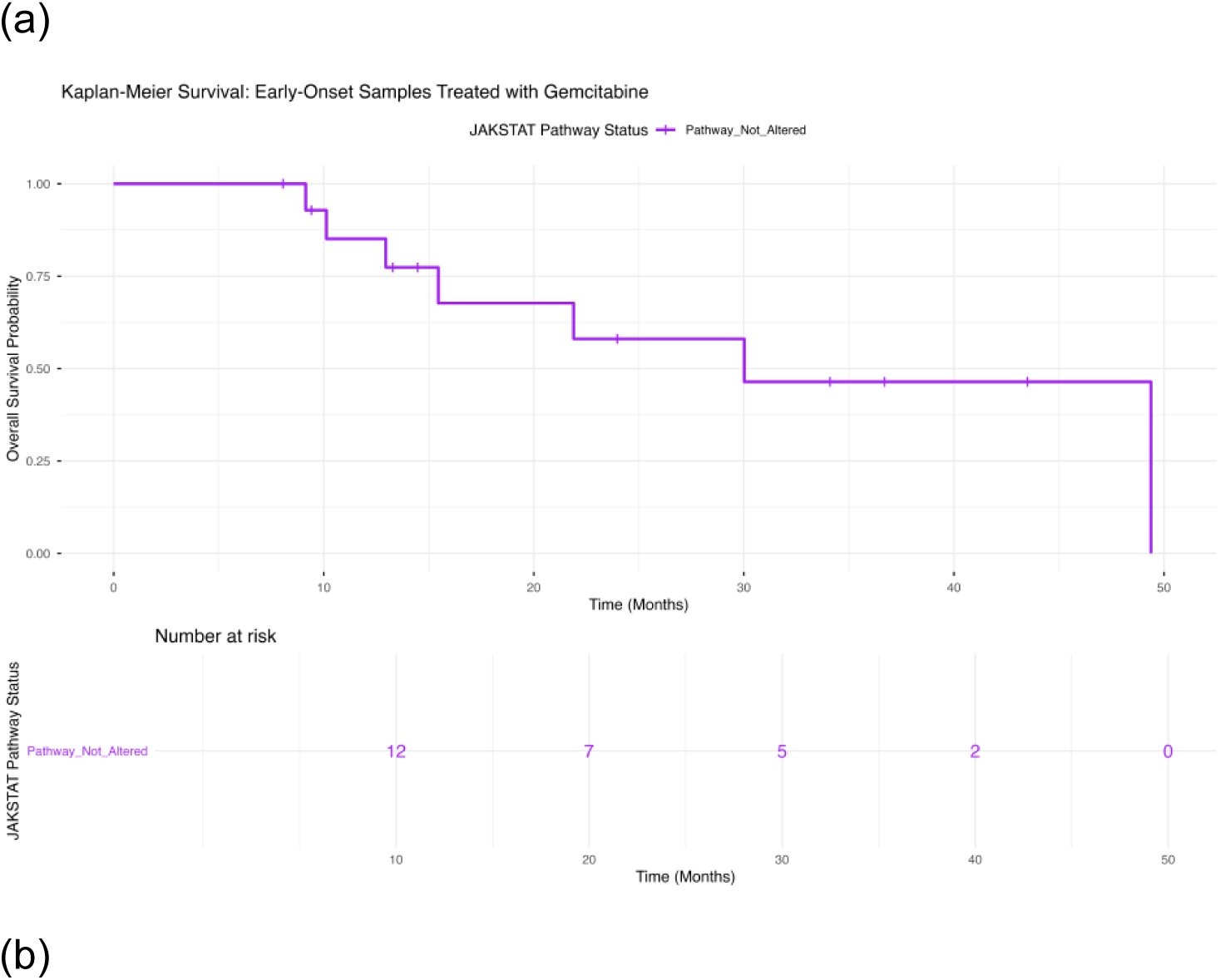
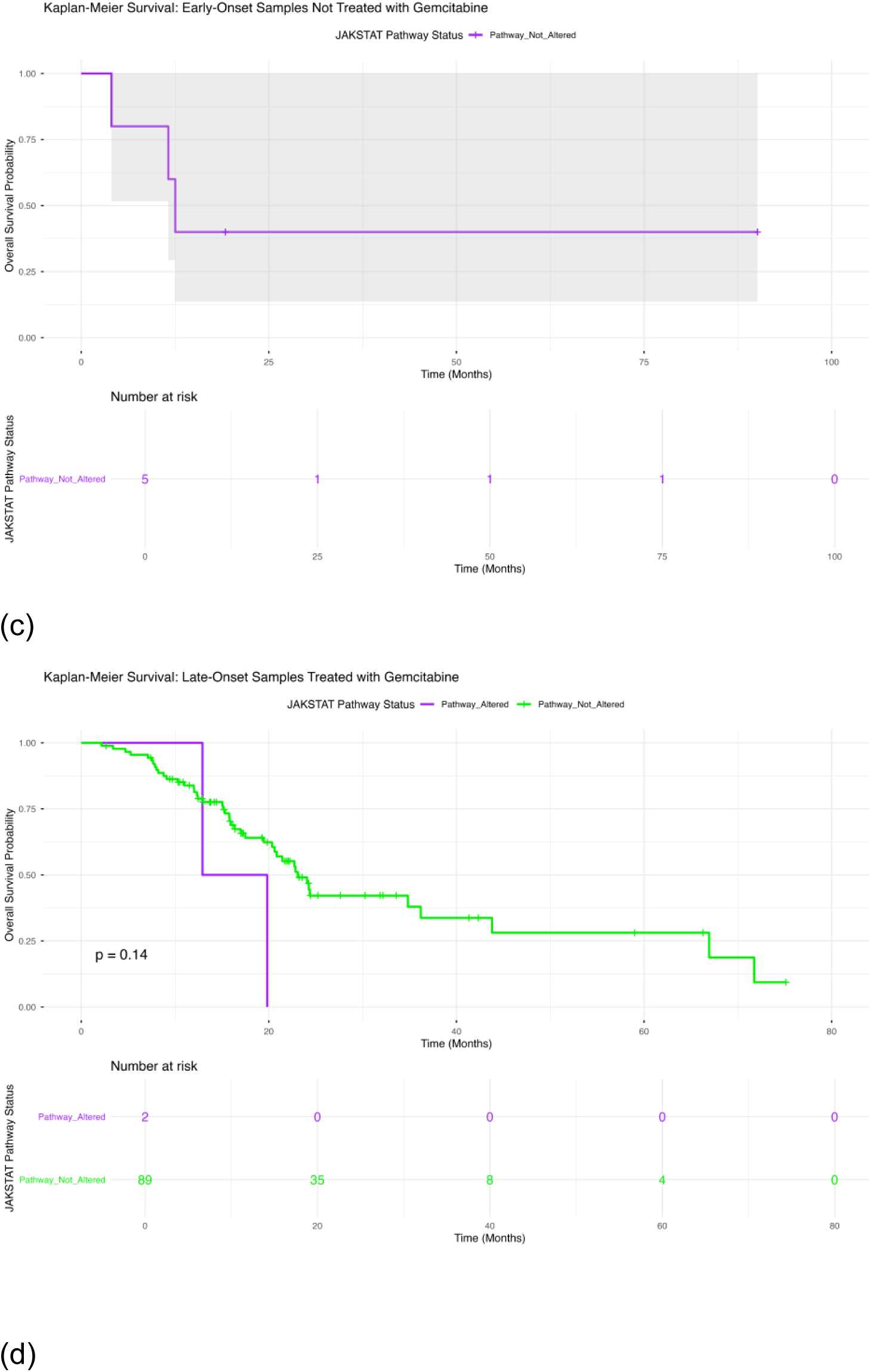
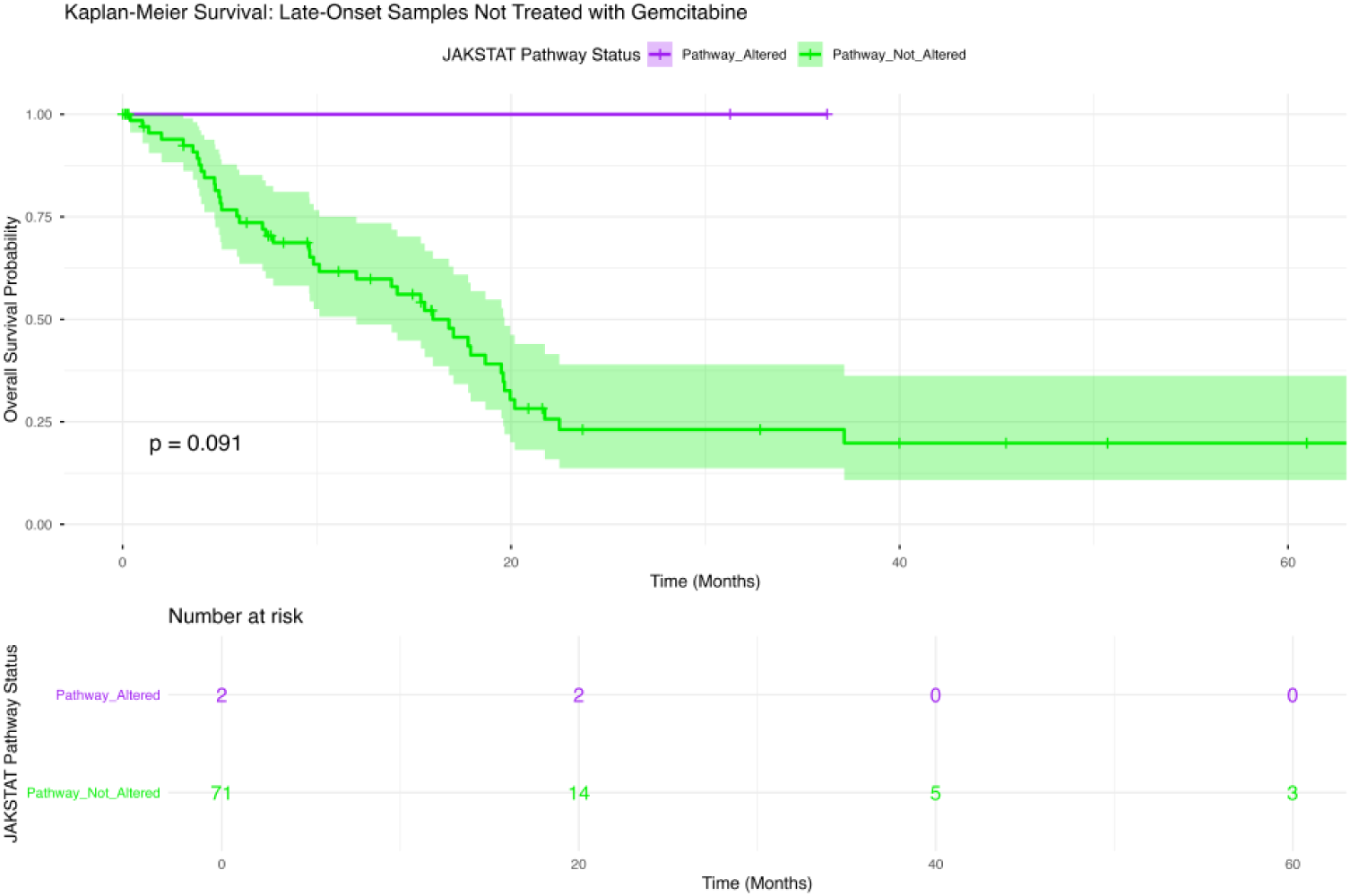
Overall survival according to JAK/STAT pathway alteration status across age- and treatment-stratified PDAC subgroups. Kaplan–Meier survival analyses compare overall survival between patients harboring JAK/STAT pathway alterations and those without detectable JAK/STAT alterations, stratified by age at diagnosis and exposure to gemcitabine-based therapy. The four panels correspond to: (a) early-onset PDAC (<50 years) treated with gemcitabine, (b) early-onset PDAC (<50 years) without gemcitabine treatment, (c) late-onset PDAC (≥50 years) treated with gemcitabine, and (d) late-onset PDAC (≥50 years) not exposed to gemcitabine. Survival patterns within each subgroup were evaluated to determine the prognostic relevance of JAK/STAT pathway dysregulation in distinct clinical contexts. Shaded areas represent 95% confidence intervals, and the accompanying risk tables display the number of patients remaining at risk at successive time points throughout follow-up.

#### 2.5.1 Early-Onset PDAC with Gemcitabine Exposure

Among early-onset PDAC patients treated with gemcitabine (Figure 2a), assessment of the prognostic significance of JAK/STAT pathway alterations was limited by the absence of tumors harboring detectable pathway alterations within this subgroup.

Consequently, survival estimates were generated exclusively from patients without JAK/STAT pathway alterations, preventing direct comparison between molecularly defined groups. The observed survival pattern demonstrated a gradual decline in overall survival probability over time, with approximately 50% of patients remaining alive beyond 30 months of follow-up. Given the lack of a comparison cohort and the relatively small number of patients at risk, these findings should be interpreted cautiously and do not permit definitive conclusions regarding the impact of JAK/STAT pathway dysregulation on survival among gemcitabine-treated early-onset PDAC patients.

#### 2.5.2 Early-Onset PDAC without Gemcitabine Exposure

Among early-onset PDAC patients who did not receive gemcitabine therapy (Figure 2b), evaluation of the prognostic relevance of JAK/STAT pathway alterations was not feasible because no tumors with detectable JAK/STAT pathway alterations were identified in this subgroup. As a result, the Kaplan–Meier survival curve represents only patients lacking JAK/STAT pathway alterations, precluding direct comparison between altered and non-altered tumors. The observed survival pattern showed an early decline in survival probability followed by a prolonged plateau during follow-up. Given the absence of a molecular comparison group and the limited sample size, no conclusions can be drawn regarding the impact of JAK/STAT pathway dysregulation on survival outcomes in untreated early-onset PDAC patients.

#### 2.5.3 Late-Onset PDAC with Gemcitabine Exposure

Among late-onset PDAC patients receiving gemcitabine-based therapy (Figure 2c), JAK/STAT pathway alteration status was not significantly associated with overall survival (p = 0.14). Patients harboring JAK/STAT pathway alterations exhibited a more rapid decline in survival during the early follow-up period compared with those lacking detectable alterations. However, the interpretation of this trend is limited by the very small number of altered cases, with only two patients represented in the JAK/STAT-altered subgroup. Although the survival curves suggest a potential adverse effect of JAK/STAT pathway alterations, the absence of statistical significance and the limited sample size preclude definitive conclusions. Larger cohorts will be necessary to clarify the prognostic role of JAK/STAT pathway dysregulation in gemcitabine-treated late-onset PDAC.

#### 2.5.4 Late-Onset PDAC without Gemcitabine Exposure

Among late-onset PDAC patients who did not receive gemcitabine therapy (Figure 2d), JAK/STAT pathway alteration status demonstrated a trend toward an association with overall survival, although statistical significance was not reached (p = 0.09). Patients harboring JAK/STAT pathway alterations appeared to experience more favorable survival outcomes compared with those lacking detectable alterations, with the survival curves separating early during follow-up and remaining distinct over time. Notably, the JAK/STAT-altered subgroup maintained a high survival probability throughout the observation period, whereas the non-altered group exhibited a progressive decline in survival. However, these findings should be interpreted cautiously due to the very limited number of patients with JAK/STAT pathway alterations, which restricts statistical power and may influence the stability of survival estimates. Nonetheless, the observed pattern suggests that JAK/STAT pathway alterations may warrant further investigation as potential prognostic biomarkers in untreated late-onset PDAC.

### 2.6 Conversational Artificial Intelligence Reveals Treatment- and Pathway-Specific Molecular Relationships in PDAC

#### 2.6.1 AI-HOPE-TGFβ: Identification of KRAS-Associated and Pathway-Level Signatures

To further investigate the biological context of TGFβ pathway dysregulation, we employed the AI-HOPE-TGFβ conversational artificial intelligence framework to perform dynamic cohort construction, enrichment testing, and large-scale association analyses within the integrated PDAC dataset. This approach enabled real-time interrogation of molecular relationships using natural language–driven clinical research questions.

As an initial exploratory analysis, the AI agent evaluated whether KRAS mutations were preferentially enriched among tumors harboring TGFβ pathway alterations (Figure S2). Through automated cohort generation, the framework identified 50 tumors with TGFβ pathway alterations and 134 tumors lacking detectable pathway abnormalities. Odds ratio analysis demonstrated a significant association between KRAS mutation status and TGFβ pathway dysregulation (odds ratio = 4.13, 95% CI: 1.80–9.47; p = 0.001), indicating that TGFβ pathway-altered tumors were substantially more likely to harbor KRAS mutations. These findings support extensive biological interaction between KRAS-driven oncogenic signaling and TGFβ-mediated tumor progression in PDAC.

To further characterize the molecular architecture of TGFβ pathway-altered tumors, AI-HOPE-TGFβ performed a comprehensive clinicogenomic association analysis (Figure S3). The strongest associations were observed with canonical TGFβ signaling components, including SMAD4, TGFBR2, and TGFBR1, confirming the biological validity of the pathway definition. In addition, significant enrichment of RTK-RAS and MAPK pathway alterations, together with KRAS mutations and RNF43 alterations, suggested coordinated activation of multiple oncogenic signaling networks. Associations with BMP-family genes, including BMP1, BMPR1A, and BMP5, further highlighted perturbation of developmental signaling pathways closely linked to TGFβ biology.

Collectively, these analyses indicate that TGFβ pathway-altered PDAC represents a distinct molecular subset characterized by extensive integration of KRAS-driven signaling, growth factor receptor pathways, and developmental regulatory networks. Importantly, the AI-HOPE-TGFβ framework enabled rapid identification of these relationships through automated statistical testing and context-aware cohort construction, demonstrating the value of conversational AI for precision oncology research.

#### 2.6.2 AI-HOPE-JAK/STAT: Discovery of Interconnected Signaling Networks and Treatment-Associated Molecular Features

We next deployed the AI-HOPE-JAK/STAT framework to investigate the genomic landscape associated with JAK/STAT pathway alterations and to evaluate potential treatment-related associations involving pathway components (Figures S5-6).

A comprehensive association analysis was first performed to identify molecular features linked to JAK/STAT pathway dysregulation (Figure S5). Although JAK/STAT pathway alterations were uncommon within the cohort, occurring in only four tumors, the AI agent identified numerous statistically significant associations involving both canonical pathway members and interconnected signaling networks. Strong relationships were observed with PIAS1, STAT1, and STAT6, confirming coherence within the JAK/STAT signaling axis. Additional associations involved receptor tyrosine kinase regulators, including NTRK1, MET, EGFR, and PDGFRA, as well as multiple components of the PI3K/AKT pathway, including PIK3R1, PIK3R2, PIK3R3, and AKT2.

Beyond these pathways, enrichment of WNT-associated genes such as CTNNB1, WIF1, AXIN2, and SFRP1 suggested extensive signaling cross-talk. Notably, significant associations with SMAD7, SMAD9, and ACVR1B revealed potential mechanistic interactions between JAK/STAT and TGFβ signaling networks, emphasizing the highly interconnected nature of PDAC biology. These findings support the concept that JAK/STAT pathway alterations define a rare but biologically distinct molecular subgroup characterized by simultaneous perturbation of multiple oncogenic pathways.

To explore potential treatment-related molecular patterns, AI-HOPE-JAK/STAT was subsequently used to evaluate whether STAT1 mutation frequency differed according to gemcitabine exposure (Figure S6). The analysis compared 106 gemcitabine-treated patients with 78 patients who had not received gemcitabine. STAT1 mutations were rare in both groups, occurring in less than 2% of tumors, and no significant association with treatment status was observed (odds ratio = 0.73, 95% CI: 0.045–11.91; p = 1.0). These findings suggest that STAT1 genomic alterations are uncommon events in PDAC and do not appear to be selectively enriched in gemcitabine-treated patients.

## 3. Discussion

The molecular determinants of therapeutic response in PDAC extend beyond individual driver mutations and increasingly involve complex interactions among signaling pathways, stromal components, immune cells, and treatment-associated selective pressures. In the present study, we employed a conversational artificial intelligence framework to investigate the genomic landscape of TGFβ and JAK/STAT signaling pathways across age- and gemcitabine-stratified PDAC cohorts. Several important observations emerged from this analysis. First, TGFβ pathway alterations were consistently detected across both early- and late-onset PDAC, regardless of gemcitabine exposure, suggesting that disruption of this pathway represents a stable feature of PDAC biology. Second, gene-level analyses revealed that this pattern was largely driven by recurrent SMAD4 alterations, whereas most other TGFβ pathway genes were altered only sporadically. Third, JAK/STAT pathway alterations were exceedingly rare throughout the cohort, with only isolated low-frequency events identified in late-onset tumors. These findings suggest that TGFβ and JAK/STAT pathways contribute to PDAC through fundamentally different genomic architectures and may represent distinct forms of biological dependency.

One of the most notable findings of this study is the remarkable stability of TGFβ pathway alteration frequencies across age groups and treatment contexts. Unlike pathways that demonstrate substantial variation according to therapeutic exposure, TGFβ pathway disruption appeared relatively conserved throughout the cohort. This observation is consistent with the central role of TGFβ signaling in PDAC pathogenesis. Previous studies have demonstrated that alterations affecting TGFβ receptors and downstream mediators occur frequently during pancreatic tumor evolution and contribute to epithelial plasticity, stromal remodeling, immune suppression, and metastatic progression. Rather than functioning solely as a treatment-associated adaptation, TGFβ pathway dysregulation may therefore represent a foundational component of PDAC biology that persists throughout disease progression.

Importantly, pathway-level analyses concealed a highly concentrated gene-level architecture. The majority of TGFβ pathway alterations were attributable to SMAD4, with relatively few mutations identified in upstream receptors or BMP-related signaling components. This finding is consistent with prior genomic studies reporting SMAD4 loss as one of the most common alterations in PDAC. The predominance of SMAD4 suggests that disruption of canonical TGFβ signal transduction remains a major mechanism through which this pathway contributes to tumor development. The comparable frequencies of SMAD4 alterations observed across age and treatment groups further support the concept that SMAD4 inactivation is an early and persistent event rather than a consequence of chemotherapy-associated selection.

The biological significance of TGFβ signaling extends beyond tumor cells themselves. Extensive experimental evidence has demonstrated that TGFβ orchestrates interactions between PDAC cells and the surrounding microenvironment, particularly cancer-associated fibroblasts (CAFs), extracellular matrix components, and immune populations. TGFβ-mediated activation of CAFs promotes fibrosis, desmoplasia, and impaired drug delivery, while simultaneously enhancing epithelial-to-mesenchymal transition, metastatic dissemination, and immune evasion. Multiple preclinical studies have shown that inhibition of TGFβ signaling can enhance gemcitabine sensitivity and reduce tumor aggressiveness, supporting the notion that this pathway remains functionally relevant even when genomic alterations are relatively stable across clinical subgroups. Consequently, the absence of treatment-associated enrichment in our genomic analysis should not be interpreted as evidence that TGFβ signaling is uninvolved in gemcitabine response. Rather, it suggests that pathway activity may be regulated through mechanisms that extend beyond DNA-level alterations, including stromal interactions, transcriptional regulation, cytokine signaling, and post-translational modifications.

In contrast, the JAK/STAT pathway demonstrated a strikingly different genomic profile. At the pathway level, alterations were detected in only a small fraction of tumors, and gene-level analyses revealed isolated mutations distributed across multiple pathway components without a dominant recurrent driver. No JAK/STAT alterations were identified in early-onset PDAC, while late-onset tumors harbored only occasional mutations affecting JAK family members, STAT transcription factors, PIAS regulators, and PTPRC. This extremely low mutational burden suggests that direct genomic disruption of JAK/STAT signaling is uncommon in PDAC.

However, the rarity of genomic alterations should not be interpreted as evidence that JAK/STAT signaling lacks biological importance. On the contrary, a growing body of literature identifies IL-6/JAK/STAT3 signaling as a central regulator of PDAC progression, immune suppression, stromal remodeling, cachexia, and therapeutic resistance. Experimental studies have demonstrated that activation of STAT3 promotes tumor cell survival, enhances fibrosis, facilitates immune escape, and contributes to resistance to gemcitabine and other cytotoxic agents. Furthermore, gemcitabine itself has been shown to induce JAK/STAT-dependent upregulation of PD-L1, highlighting the dynamic relationship between chemotherapy and immune signaling pathways. The discrepancy between the extensive functional literature and the low genomic alteration rates observed in this study likely reflects the fact that JAK/STAT pathway activation is predominantly driven through cytokine-mediated signaling and microenvironmental stimuli rather than recurrent somatic mutations.

This distinction highlights an important conceptual lesson for precision oncology. Pathway activity and pathway mutation are not synonymous. While genomic analyses are highly informative for identifying pathways driven by recurrent structural alterations, they may underestimate the biological relevance of signaling networks that are activated primarily through transcriptional, inflammatory, or stromal mechanisms. The TGFβ and JAK/STAT pathways provide complementary examples of this principle. TGFβ signaling exhibits recurrent genomic disruption, primarily through SMAD4 alterations, whereas JAK/STAT signaling appears to be largely regulated through non-genomic mechanisms. Consequently, integrated analyses incorporating transcriptomic, phosphoproteomic, spatial, and immune microenvironment data will likely be required to fully characterize the clinical significance of JAK/STAT activity in PDAC.

An additional contribution of this study is methodological. The AI-HOPE framework enabled rapid interrogation of complex clinicogenomic datasets through natural language-driven cohort construction, pathway analysis, and gene-level exploration. Precision oncology increasingly requires iterative analyses across multiple overlapping clinical variables, including age, treatment exposure, molecular subtype, and survival outcome. Traditional approaches often require repeated coding, data restructuring, and manual query generation. Conversational artificial intelligence provides a complementary analytical paradigm that allows investigators to rapidly formulate, refine, and validate hypotheses while maintaining reproducibility through conventional statistical testing. The present study demonstrates how AI-assisted interrogation can facilitate discovery of biologically meaningful patterns that may otherwise remain obscured within large multidimensional datasets.

Several limitations should be considered when interpreting these findings. First, the early-onset PDAC subgroup was relatively small, limiting statistical power and increasing the possibility of type II error. Second, gemcitabine exposure was analyzed as a binary variable and did not capture differences in treatment duration, dose intensity, combination regimens, or timing relative to molecular profiling. Third, the cohort consisted predominantly of stage II tumors, which may limit extrapolation to metastatic disease. Fourth, genomic alterations do not necessarily correspond to functional pathway activation, particularly for signaling networks such as JAK/STAT that are strongly influenced by extracellular cytokines and tumor-stroma interactions. Finally, the study focused on DNA-level alterations and therefore could not evaluate transcriptional, epigenetic, proteomic, or spatial determinants of pathway activity.

Future studies should expand upon these findings through integration of multi-omic datasets capable of capturing pathway activity beyond genomic alterations alone. Transcriptomic and phosphoproteomic profiling may be particularly informative for evaluating JAK/STAT signaling, while spatial analyses could clarify how TGFβ-driven stromal remodeling influences therapeutic response. Larger cohorts with detailed treatment annotation and longitudinal sampling will also be necessary to determine whether specific pathway states evolve during chemotherapy exposure. Functional studies examining the interaction between SMAD4 status, stromal composition, and gemcitabine sensitivity may further clarify the translational implications of TGFβ pathway disruption.

In summary, this study demonstrates that TGFβ and JAK/STAT signaling pathways exhibit markedly different genomic architectures in PDAC. TGFβ pathway alterations are relatively common and largely driven by recurrent SMAD4 disruption, supporting a stable role for this pathway in pancreatic tumor biology. In contrast, JAK/STAT pathway alterations are rare and distributed across multiple low-frequency genes, suggesting that pathway activation is primarily mediated through non-genomic mechanisms. These findings underscore the importance of distinguishing pathway mutation from pathway activity and highlight the value of conversational artificial intelligence as a scalable framework for precision oncology research. Together, these results support continued investigation of TGFβ- and JAK/STAT-centered therapeutic strategies while demonstrating how AI-enabled analyses can accelerate biologically informed hypothesis generation in complex clinical-genomic datasets.

## 4. Materials and Methods

### Cohort Assembly and Clinical Data Collection

This retrospective study analyzed a cohort of 184 patients diagnosed with PDAC who underwent comprehensive molecular profiling. Clinical and genomic information was obtained from harmonized clinicogenomic datasets containing patient demographics, tumor characteristics, treatment histories, and survival outcomes. Only primary tumor specimens were included to minimize biological variability associated with metastatic disease and treatment-induced molecular evolution.

Patients were categorized according to age at diagnosis and exposure to gemcitabine-containing regimens. Early-onset PDAC was defined as diagnosis before 50 years of age, whereas patients diagnosed at or after 50 years were classified as late-onset PDAC. Treatment groups were established based on documented administration of gemcitabine during the clinical course. These classifications generated four clinically relevant subgroups: early-onset gemcitabine-treated, early-onset non-gemcitabine-treated, late-onset gemcitabine-treated, and late-onset non-gemcitabine-treated PDAC.

To ensure analytical consistency, duplicate patient entries and repeated molecular profiles were removed. When multiple sequencing records were available for a single patient, the sample containing the most complete genomic information was retained for downstream analyses.

### TGFβ and JAK/STAT Pathway Characterization

Two biologically relevant signaling networks were selected for investigation: the transforming growth factor-beta (TGFβ) pathway and the Janus kinase/signal transducer and activator of transcription (JAK/STAT) pathway. These pathways were chosen because of their established roles in pancreatic tumor initiation, immune regulation, metastatic progression, therapeutic resistance, and modulation of the tumor microenvironment.

Pathway-specific gene sets were curated using established molecular pathway databases and published literature. The TGFβ pathway panel included core ligands, receptors, intracellular mediators, and downstream transcriptional regulators involved in canonical TGFβ signaling. The JAK/STAT pathway panel consisted of cytokine receptors, JAK family kinases, STAT transcription factors, and associated regulatory components responsible for signal transduction and immune-related cellular responses.

Somatic genomic alterations were extracted from next-generation sequencing datasets. Only protein-altering variants, including missense mutations, nonsense mutations, frameshift insertions/deletions, splice-site variants, and translation start-site disruptions, were considered. Synonymous variants and alterations lacking predicted functional consequences were excluded. A tumor was classified as pathway-altered when at least one qualifying mutation was identified within the corresponding pathway gene set.

### Molecular and Survival Analyses

Pathway alteration frequencies were calculated across the entire cohort and within age- and treatment-defined subgroups. Comparative analyses were performed to identify differences in the prevalence of TGFβ and JAK/STAT pathway alterations between clinically distinct patient populations.

Overall survival served as the primary clinical endpoint and was defined as the interval between diagnosis and death or last documented follow-up. Kaplan–Meier methodology was used to estimate survival distributions, and statistical differences between groups were evaluated using the log-rank test. Survival analyses were conducted separately within each age- and treatment-stratified subgroup to investigate the context-dependent prognostic significance of pathway alterations.

Associations between categorical clinical variables and pathway status were assessed using Fisher’s exact test or Pearson’s chi-square test, as appropriate. Statistical significance was defined using a two-sided p-value threshold of <0.05.

### Conversational Artificial Intelligence Framework

To facilitate rapid and reproducible interrogation of clinicogenomic relationships, analyses were performed using the Artificial Intelligence–Health Outcomes and Precision Oncology Ecosystem (AI-HOPE), a conversational artificial intelligence platform designed for precision oncology research. AI-HOPE enables investigators to interact with complex molecular and clinical datasets through natural language queries, reducing barriers to large-scale data exploration and hypothesis generation.

Specialized AI agents focused on TGFβ and JAK/STAT signaling pathways were deployed to identify pathway alterations, generate subgroup-specific mutation frequencies, construct analytical cohorts, and perform survival analyses. Through an automated query-processing framework, user-generated requests were translated into structured analytical workflows capable of executing reproducible statistical evaluations across multiple clinical strata.

### Validation and Reproducibility

To ensure methodological robustness, all outputs generated by the AI-driven framework were independently verified using conventional bioinformatics and statistical pipelines. Pathway alteration frequencies, patient subgroup assignments, and survival estimates were cross-validated to confirm concordance between AI-assisted and traditional analytical approaches. This validation strategy ensured data integrity, reproducibility, and transparency while demonstrating the utility of conversational artificial intelligence agents for precision medicine–oriented pancreatic cancer research.

## Data Availability

All data used in the present study is publicly available at https://www.cbioportal.org/ and https://genie.cbioportal.org. The datasets used in our study were aggregated/summary data, and no individual-level data were used. Additional data can be provided upon reasonable request to the authors

## Acknowledgments

The authors gratefully acknowledge Dr. Steven T. Rosen at City of Hope; the Wall Fund for Pancreatic Cancer Research at City of Hope; the Department of Integrative Translational Sciences; the Cancer Control and Population Sciences Program at the City of Hope Comprehensive Cancer Center; the Drug Development and Capacity Building: A UCR/CoH-CCC Partnership project; and the Cancer Moonshot PE-CGS initiative supported by the National Cancer Institute (NCI).

## Supplementary Materials

**Table S1.** Comparison of Early-Onset PDAC Patients Treated with Gemcitabine Versus Those Not Treated with Gemcitabine.

| TGF-beta Pathway |  |  |  |
| --- | --- | --- | --- |
| Gene | Early-Onset<br>Treated with Gemcitabine<br>n (%) | Early-Onset<br>Not Treated with Gemcitabine<br>n (%) | p-value |
| BMP1 Mutation |  |  |  |
| Present | 0 (0.0%) | 0 (0.0%) | 1 |
| Absent | 15 (100.0%) | 5 (100.0%) |  |
| BMP2K Mutation |  |  |  |
| Present | 0 (0.0%) | 0 (0.0%) | 1 |
| Absent | 15 (100.0%) | 5 (100.0%) |  |
| BMP3 Mutation |  |  |  |
| Present | 0 (0.0%) | 0 (0.0%) | 1 |
| Absent | 15 (100.0%) | 5 (100.0%) |  |
| BMP4 Mutation |  |  |  |
| Present | 0 (0.0%) | 0 (0.0%) | 1 |
| Absent | 15 (100.0%) | 5 (100.0%) |  |
| BMP5 Mutation |  |  |  |
| Present | 0 (0.0%) | 0 (0.0%) | 1 |
| Absent | 15 (100.0%) | 5 (100.0%) |  |
| BMP7 Mutation |  |  |  |
| Present | 0 (0.0%) | 0 (0.0%) | 1 |
| Absent | 15 (100.0%) | 5 (100.0%) |  |
| BMP8B Mutation |  |  |  |
| Present | 0 (0.0%) | 0 (0.0%) | 1 |
| Absent | 15 (100.0%) | 5 (100.0%) |  |
| BMPER Mutation |  |  |  |
| Present | 0 (0.0%) | 0 (0.0%) | 1 |
| Absent | 15 (100.0%) | 5 (100.0%) |  |
| BMPR1A Mutation |  |  |  |
| Present | 0 (0.0%) | 0 (0.0%) | 1 |
| Absent | 15 (100.0%) | 5 (100.0%) |  |
| BMPR2 Mutation |  |  |  |
| Present | 0 (0.0%) | 0 (0.0%) | 1 |
| Absent | 15 (100.0%) | 5 (100.0%) |  |
| SMAD2 Mutation |  |  |  |
| Present | 0 (0.0%) | 0 (0.0%) | 1 |
| Absent | 15 (100.0%) | 5 (100.0%) |  |
| SMAD4 Mutation |  |  |  |
| Present | 3 (20.0%) | 0 (0.0%) | 0.5395 |
| Absent | 12 (80.0%) | 5 (100.0%) |  |
| SMAD7 Mutation |  |  |  |
| Present | 0 (0.0%) | 0 (0.0%) | 1 |
| Absent | 15 (100.0%) | 5 (100.0%) |  |
| SMAD9 Mutation |  |  |  |
| Present | 0 (0.0%) | 0 (0.0%) | 1 |
| Absent | 15 (100.0%) | 5 (100.0%) |  |
| TGFB1 Mutation |  |  |  |
| Present | 0 (0.0%) | 0 (0.0%) | 1 |
| Absent | 15 (100.0%) | 5 (100.0%) |  |
| TGFBI Mutation |  |  |  |
| Present | 0 (0.0%) | 0 (0.0%) | 1 |
| Absent | 15 (100.0%) | 5 (100.0%) |  |
| TGFBR1 Mutation |  |  |  |
| Present | 0 (0.0%) | 0 (0.0%) | 1 |
| Absent | 15 (100.0%) | 5 (100.0%) |  |
| TGFBR2 Mutation |  |  |  |
| Present | 1 (6.7%) | 0 (0.0%) | 1 |
| Absent | 14 (93.3%) | 5 (100.0%) |  |
| TGFBRAP1Mutation |  |  |  |
| Present | 0 (0.0%) | 0 (0.0%) | 1 |
| Absent | 15 (100.0%) | 5 (100.0%) |  |

**Table S2.** Comparison of Late-Onset PDAC Patients Treated with Gemcitabine Versus Those Not Treated with Gemcitabine.

| TGF-beta Pathway |  |  |  |
| --- | --- | --- | --- |
| Gene | Early-Onset<br>Treated with Gemcitabine<br>n (%) | Early-Onset<br>Not Treated with Gemcitabine<br>n (%) | p-value |
| BMP1 Mutation |  |  |  |
| Present | 0 (0.0%) | 0 (0.0%) | 1 |
| Absent | 15 (100.0%) | 5 (100.0%) |  |
| BMP2K Mutation |  |  |  |
| Present | 0 (0.0%) | 0 (0.0%) | 1 |
| Absent | 15 (100.0%) | 5 (100.0%) |  |
| BMP3 Mutation |  |  |  |
| Present | 0 (0.0%) | 0 (0.0%) | 1 |
| Absent | 15 (100.0%) | 5 (100.0%) |  |
| BMP4 Mutation |  |  |  |
| Present | 0 (0.0%) | 0 (0.0%) | 1 |
| Absent | 15 (100.0%) | 5 (100.0%) |  |
| BMP5 Mutation |  |  |  |
| Present | 0 (0.0%) | 0 (0.0%) | 1 |
| Absent | 15 (100.0%) | 5 (100.0%) |  |
| BMP7 Mutation |  |  |  |
| Present | 0 (0.0%) | 0 (0.0%) | 1 |
| Absent | 15 (100.0%) | 5 (100.0%) |  |
| BMP8B Mutation |  |  |  |
| Present | 0 (0.0%) | 0 (0.0%) | 1 |
| Absent | 15 (100.0%) | 5 (100.0%) |  |
| BMPER Mutation |  |  |  |
| Present | 0 (0.0%) | 0 (0.0%) | 1 |
| Absent | 15 (100.0%) | 5 (100.0%) |  |
| BMPR1A Mutation |  |  |  |
| Present | 0 (0.0%) | 0 (0.0%) | 1 |
| Absent | 15 (100.0%) | 5 (100.0%) |  |
| BMPR2 Mutation |  |  |  |
| Present | 0 (0.0%) | 0 (0.0%) | 1 |
| Absent | 15 (100.0%) | 5 (100.0%) |  |
| **SMAD2 Mutation** |  |  |  |
| Present | 0 (0.0%) | 0 (0.0%) | 1 |
| Absent | 15 (100.0%) | 5 (100.0%) |  |
| **SMAD4 Mutation** |  |  |  |
| Present | 3 (20.0%) | 0 (0.0%) | 0.5395 |
| Absent | 12 (80.0%) | 5 (100.0%) |  |
| **SMAD7 Mutation** |  |  |  |
| Present | 0 (0.0%) | 0 (0.0%) | 1 |
| Absent | 15 (100.0%) | 5 (100.0%) |  |
| **SMAD9 Mutation** |  |  |  |
| Present | 0 (0.0%) | 0 (0.0%) | 1 |
| Absent | 15 (100.0%) | 5 (100.0%) |  |
| **TGFB1 Mutation** |  |  |  |
| Present | 0 (0.0%) | 0 (0.0%) | 1 |
| Absent | 15 (100.0%) | 5 (100.0%) |  |
| **TGFBI Mutation** |  |  |  |
| Present | 0 (0.0%) | 0 (0.0%) | 1 |
| Absent | 15 (100.0%) | 5 (100.0%) |  |
| **TGFBR1 Mutation** |  |  |  |
| Present | 0 (0.0%) | 0 (0.0%) | 1 |
| Absent | 15 (100.0%) | 5 (100.0%) |  |
| **TGFBR2 Mutation** |  |  |  |
| Present | 1 (6.7%) | 0 (0.0%) | 1 |
| Absent | 14 (93.3%) | 5 (100.0%) |  |
| **TGFBRAP1Mutation** |  |  |  |
| Present | 0 (0.0%) | 0 (0.0%) | 1 |
| Absent | 15 (100.0%) | 5 (100.0%) |  |

**Table S3.**
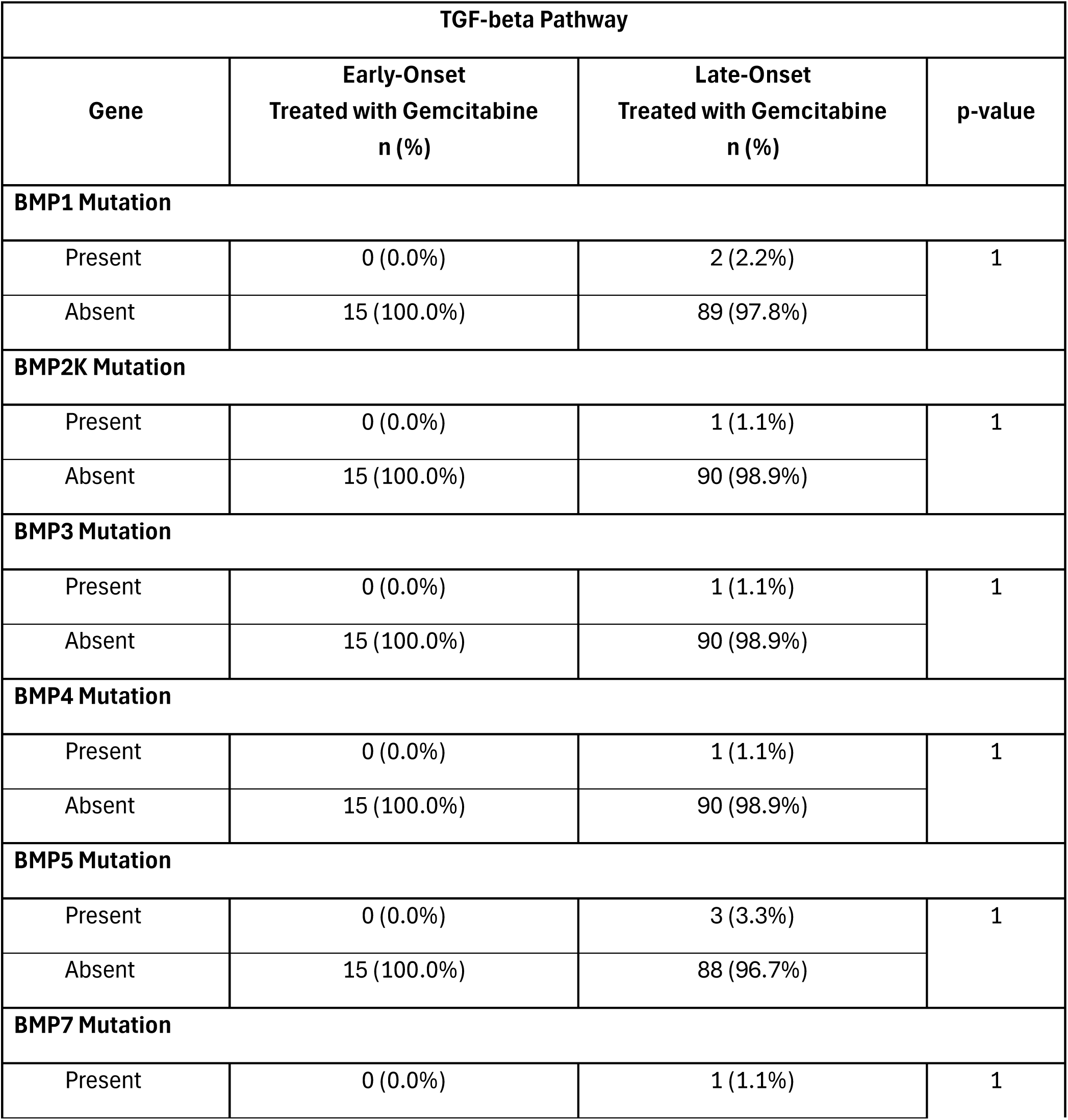
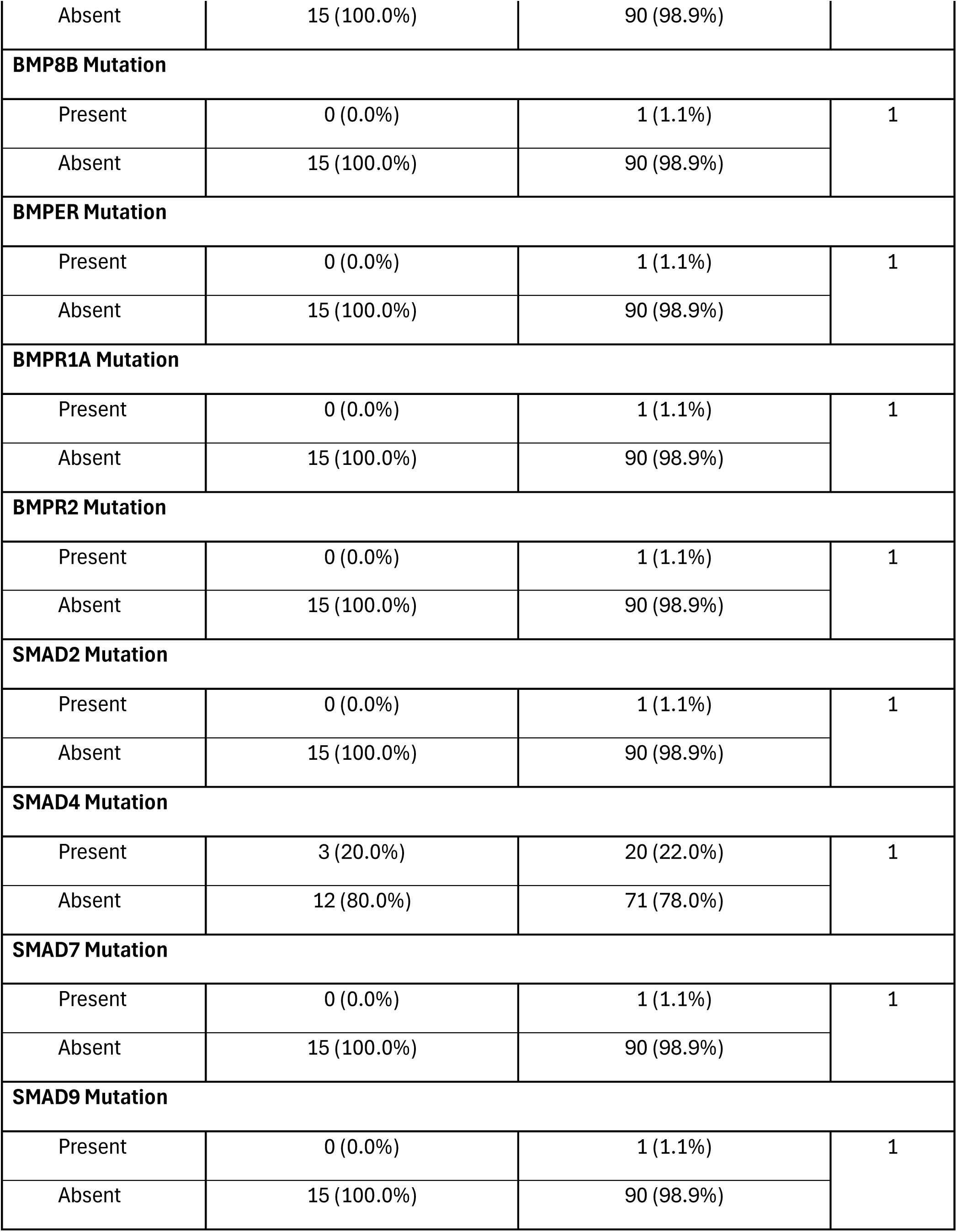
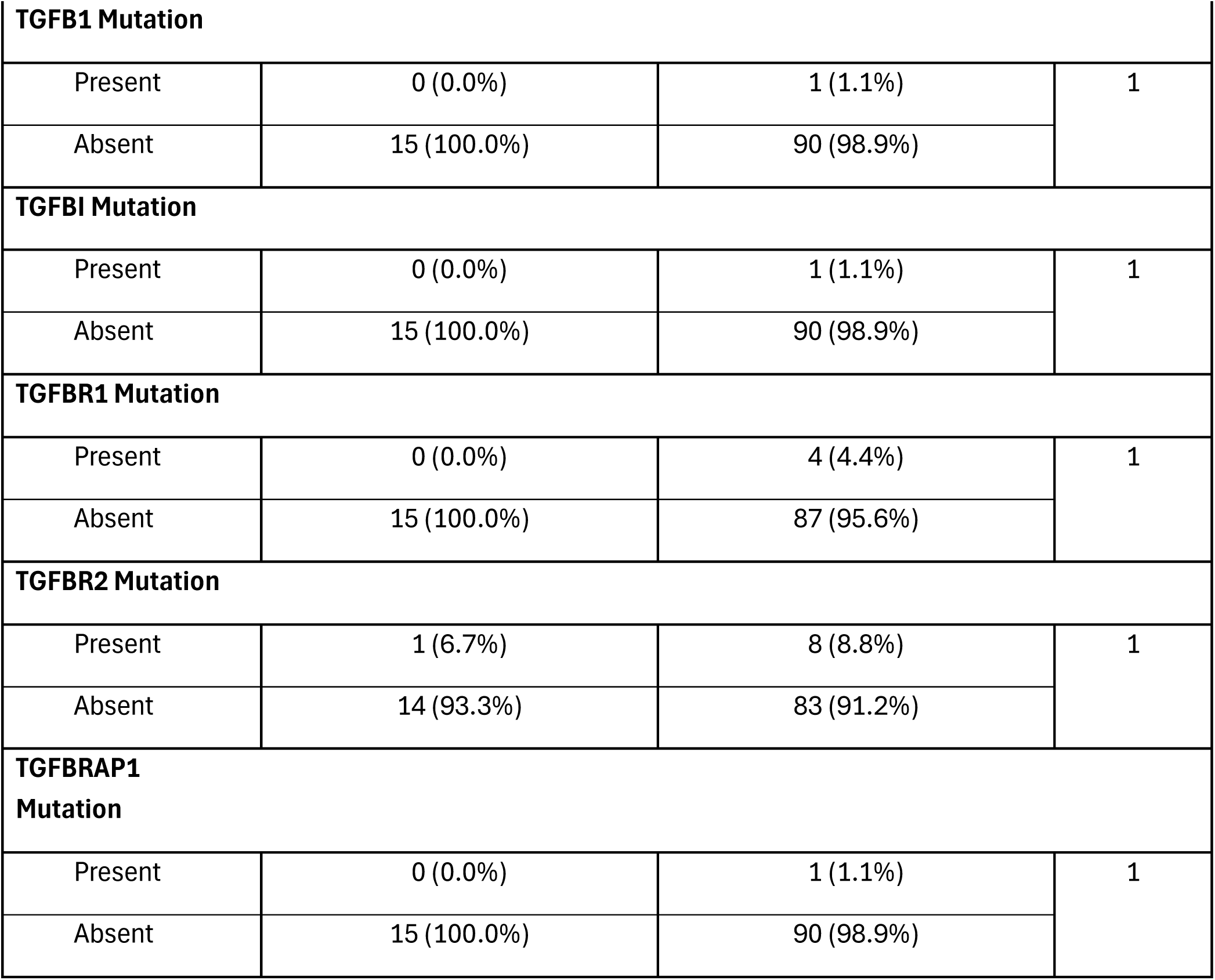
Comparison of Early-Onset PDAC Patients Versus Late-Onset PDAC Patients Treated with Gemcitabine.

**Table S4.**
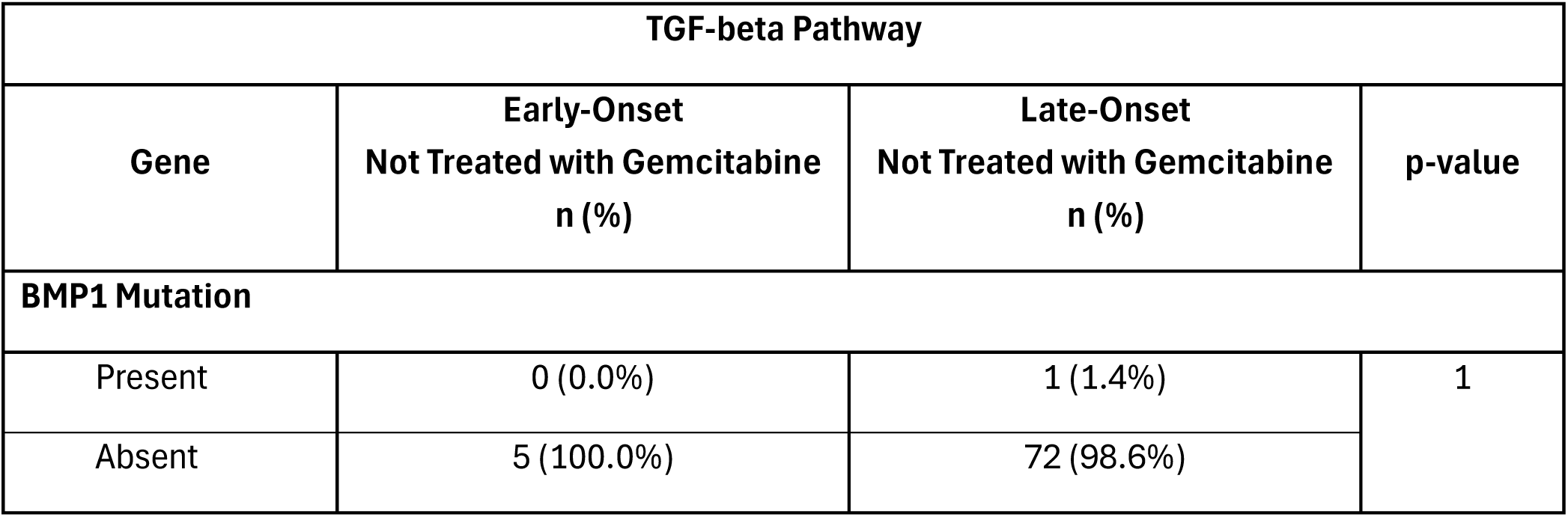
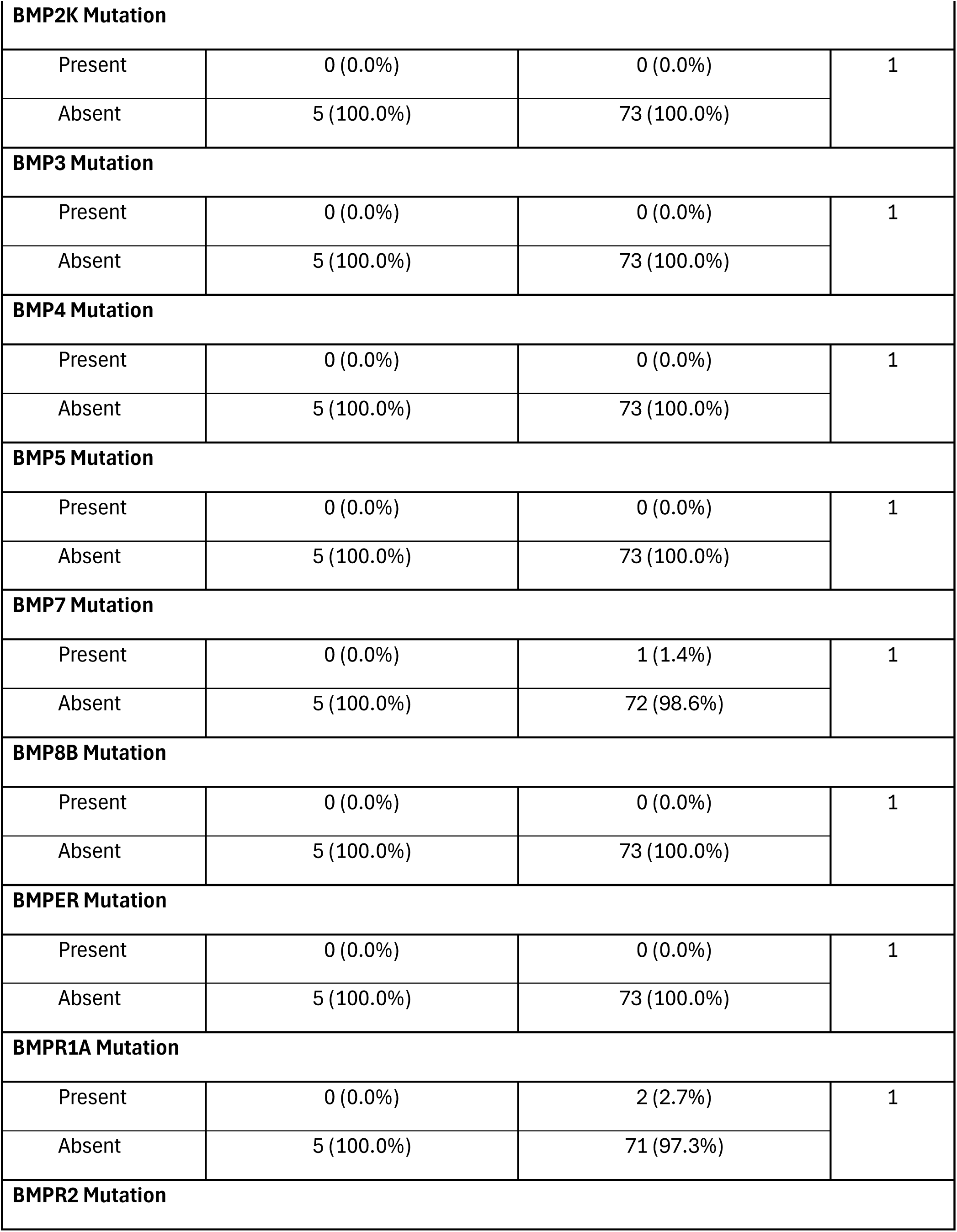
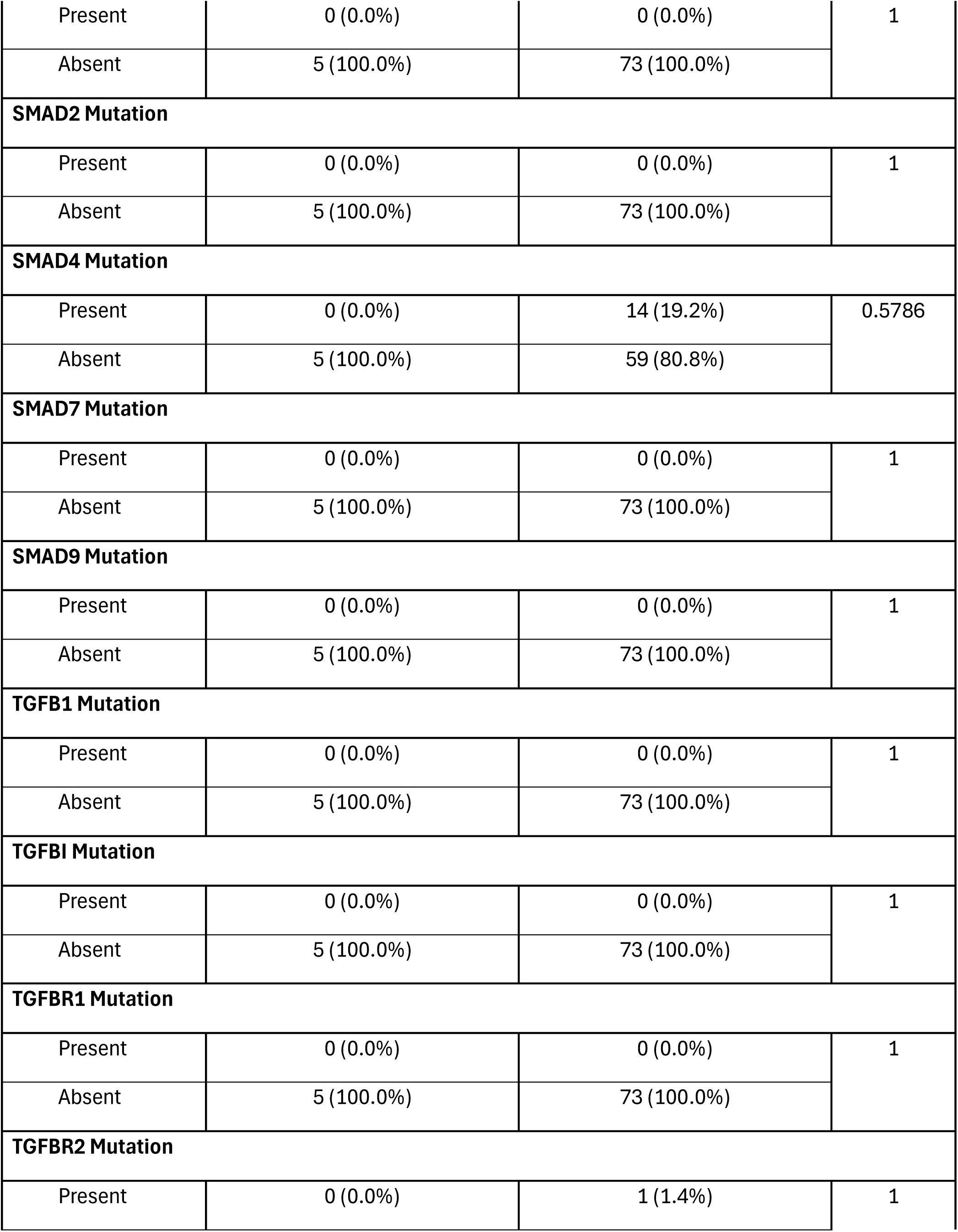
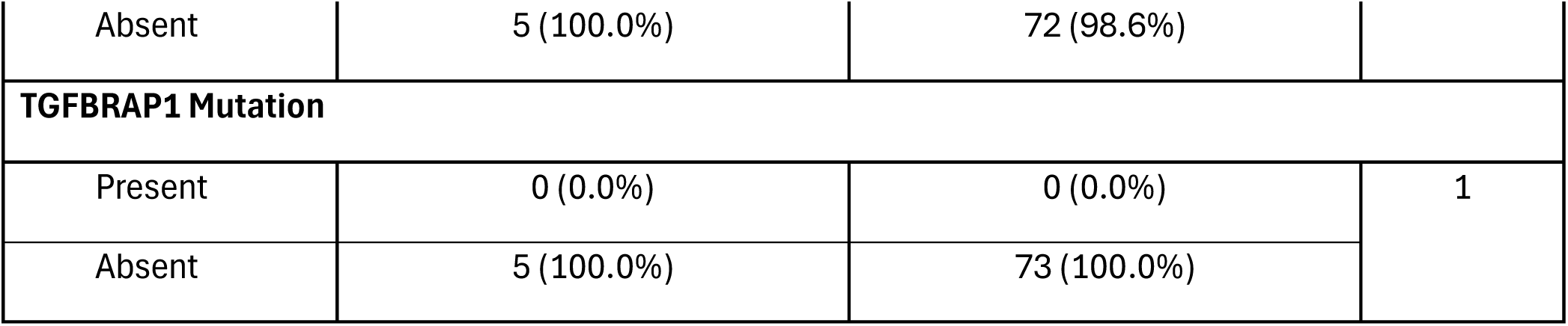
Comparison of Early-Onset PDAC Patients Versus Late-Onset PDAC Patients Not Treated with Gemcitabine.

**Table S5.**
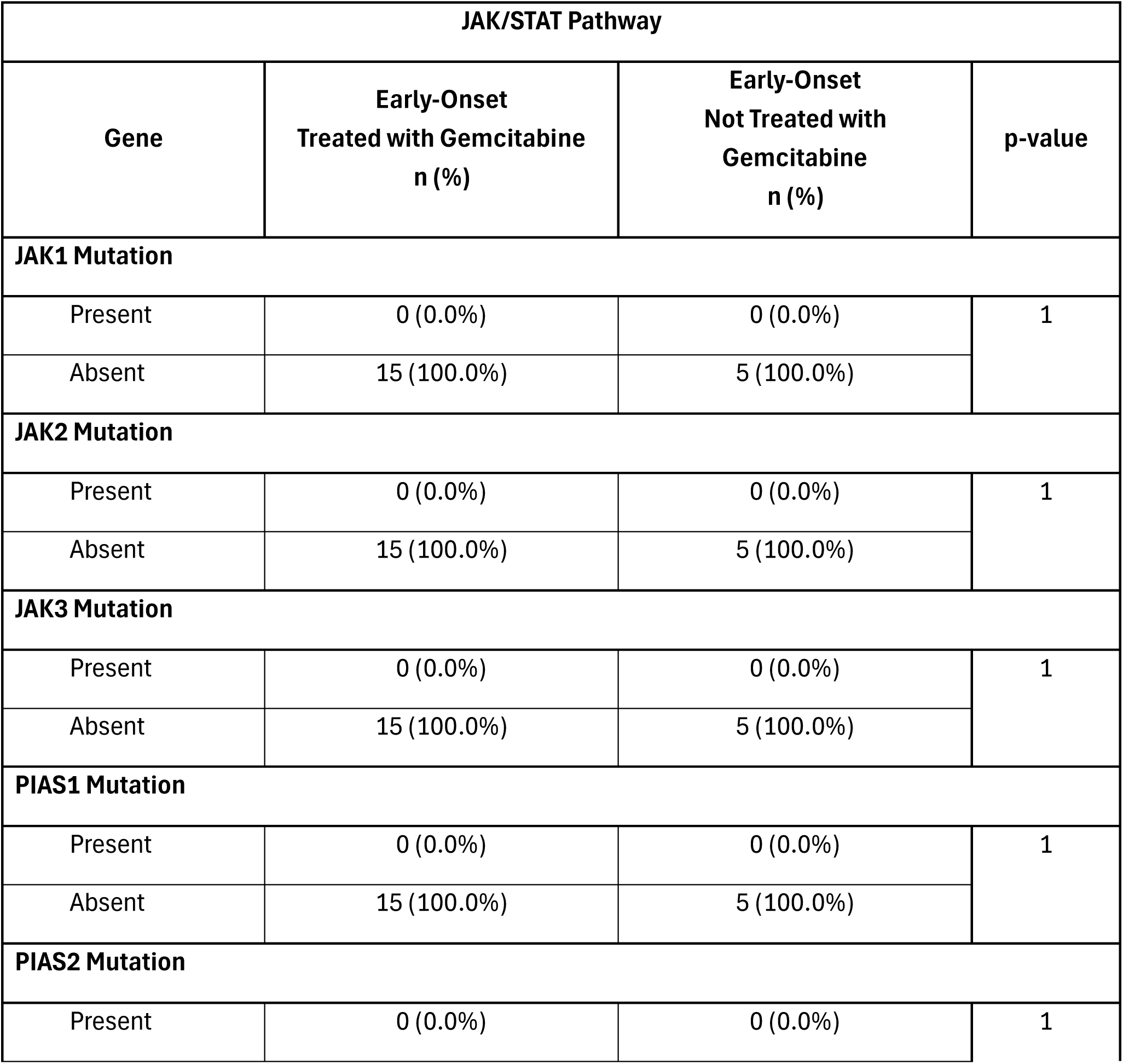
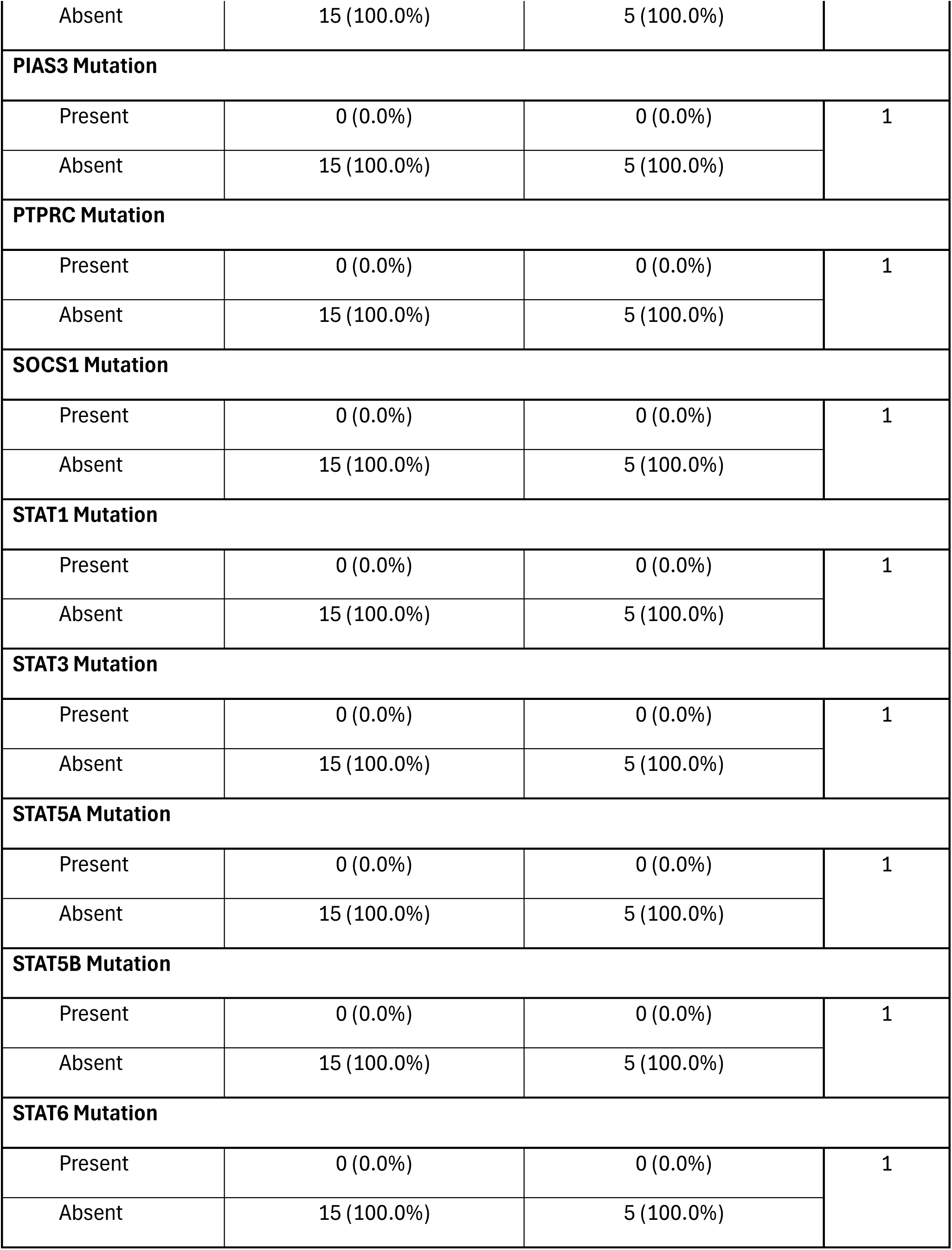
Comparison of Early-Onset PDAC Patients Treated with Gemcitabine Versus Those Not Treated with Gemcitabine.

**Table S6.**
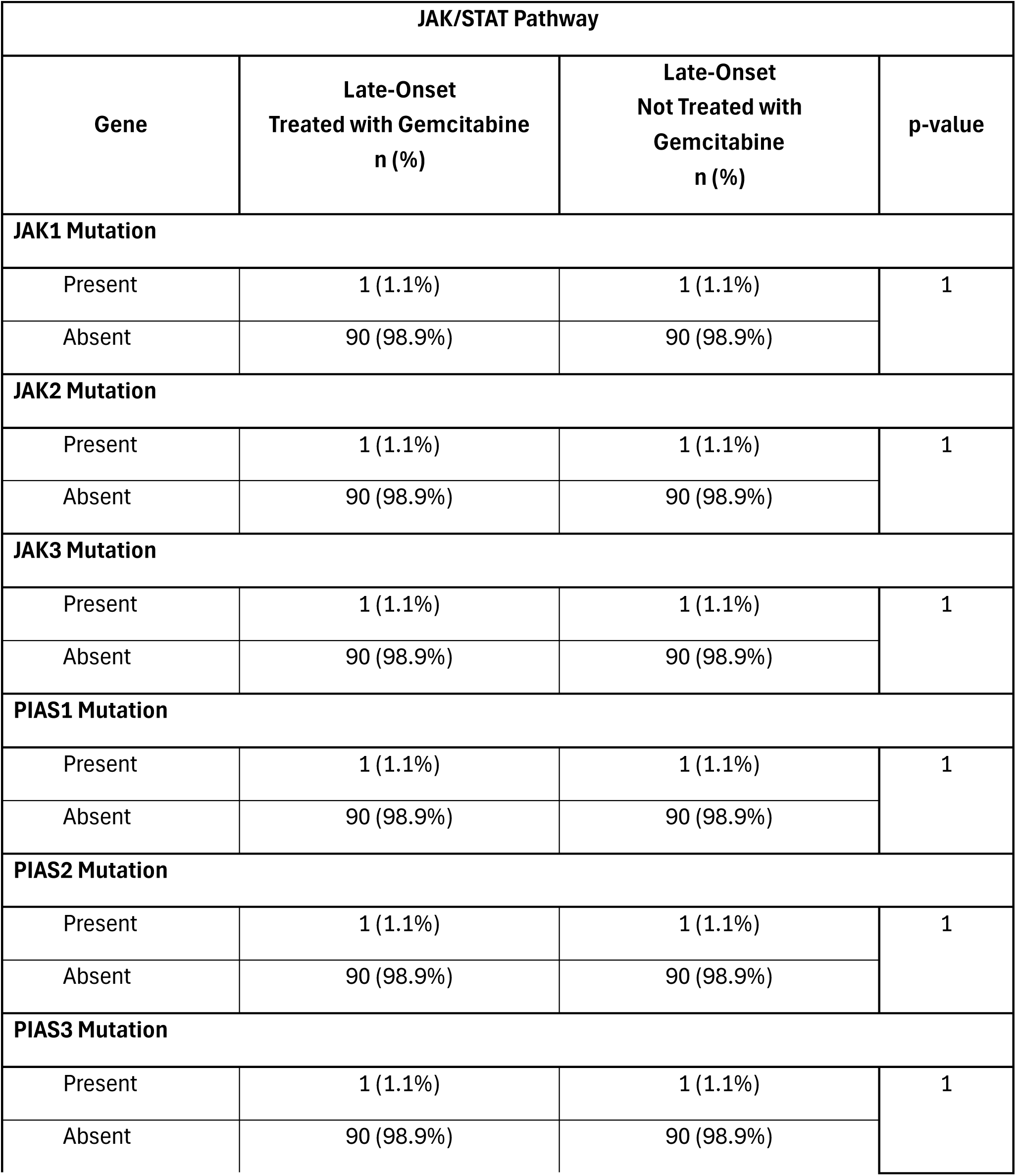
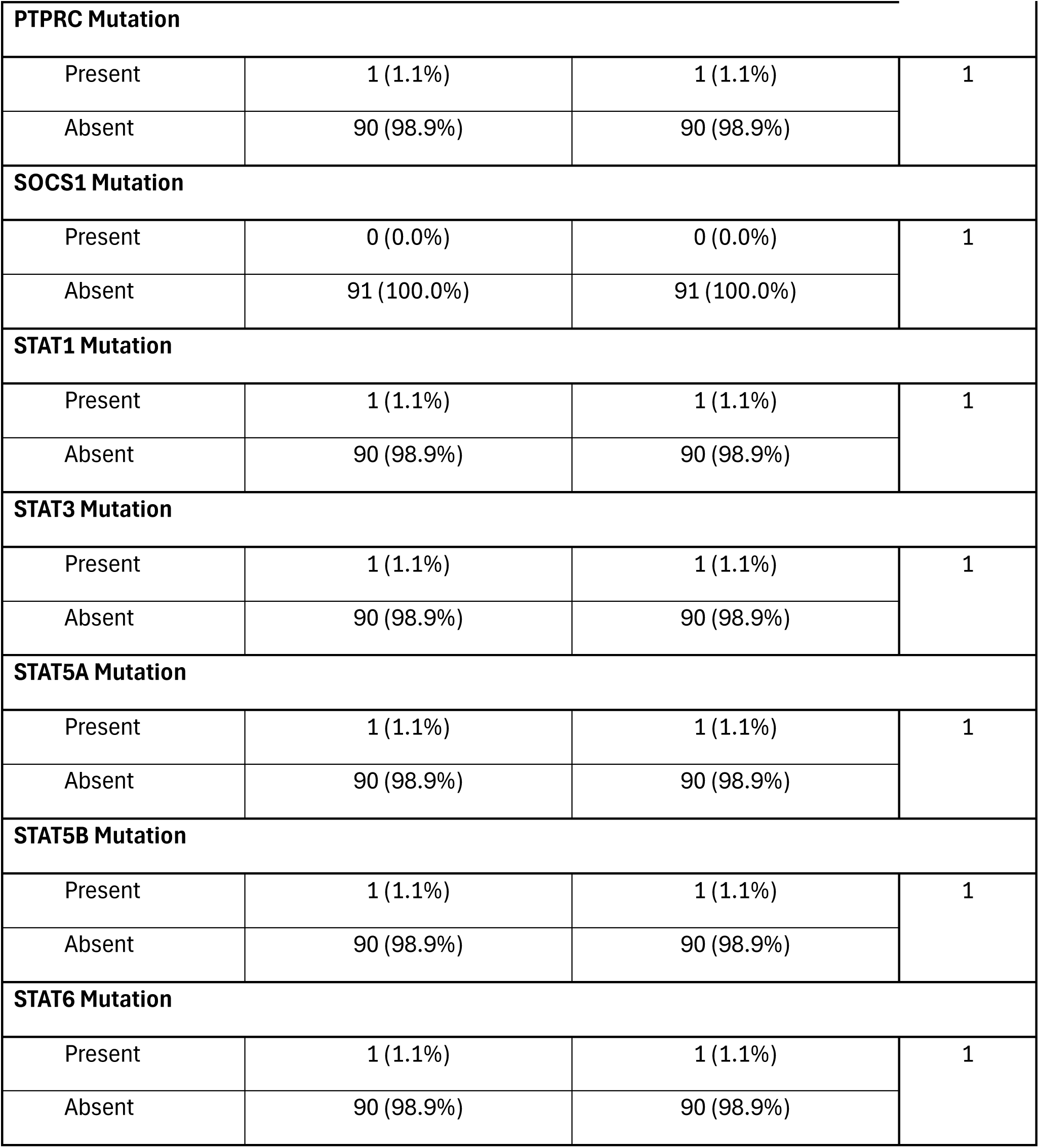
Comparison of Late-Onset PDAC Patients Treated with Gemcitabine Versus Those Not Treated with Gemcitabine.

| JAK/STAT Pathway |  |  |  |
| --- | --- | --- | --- |
| Gene | Late-Onset<br>Treated with Gemcitabine<br>n (%) | Late-Onset<br>Not Treated with<br>Gemcitabine<br>n (%) | p-value |
| JAK1 Mutation |  |  |  |
| Present | 1 (1.1%) | 1 (1.1%) | 1 |
| Absent | 90 (98.9%) | 90 (98.9%) |  |
| JAK2 Mutation |  |  |  |
| Present | 1 (1.1%) | 1 (1.1%) | 1 |
| Absent | 90 (98.9%) | 90 (98.9%) |  |
| JAK3 Mutation |  |  |  |
| Present | 1 (1.1%) | 1 (1.1%) | 1 |
| Absent | 90 (98.9%) | 90 (98.9%) |  |
| PIAS1 Mutation |  |  |  |
| Present | 1 (1.1%) | 1 (1.1%) | 1 |
| Absent | 90 (98.9%) | 90 (98.9%) |  |
| PIAS2 Mutation |  |  |  |
| Present | 1 (1.1%) | 1 (1.1%) | 1 |
| Absent | 90 (98.9%) | 90 (98.9%) |  |
| PIAS3 Mutation |  |  |  |
| Present | 1 (1.1%) | 1 (1.1%) | 1 |
| Absent | 90 (98.9%) | 90 (98.9%) |  |

| PTPRC Mutation |  |  |  |
| --- | --- | --- | --- |
| Present | 1 (1.1%) | 1 (1.1%) | 1 |
| Absent | 90 (98.9%) | 90 (98.9%) |  |
| SOCS1 Mutation |  |  |  |
| Present | 0 (0.0%) | 0 (0.0%) | 1 |
| Absent | 91 (100.0%) | 91 (100.0%) |  |
| STAT1 Mutation |  |  |  |
| Present | 1 (1.1%) | 1 (1.1%) | 1 |
| Absent | 90 (98.9%) | 90 (98.9%) |  |
| STAT3 Mutation |  |  |  |
| Present | 1 (1.1%) | 1 (1.1%) | 1 |
| Absent | 90 (98.9%) | 90 (98.9%) |  |
| STAT5A Mutation |  |  |  |
| Present | 1 (1.1%) | 1 (1.1%) | 1 |
| Absent | 90 (98.9%) | 90 (98.9%) |  |
| STAT5B Mutation |  |  |  |
| Present | 1 (1.1%) | 1 (1.1%) | 1 |
| Absent | 90 (98.9%) | 90 (98.9%) |  |
| STAT6 Mutation |  |  |  |
| Present | 1 (1.1%) | 1 (1.1%) | 1 |
| Absent | 90 (98.9%) | 90 (98.9%) |  |

**Table S7.**
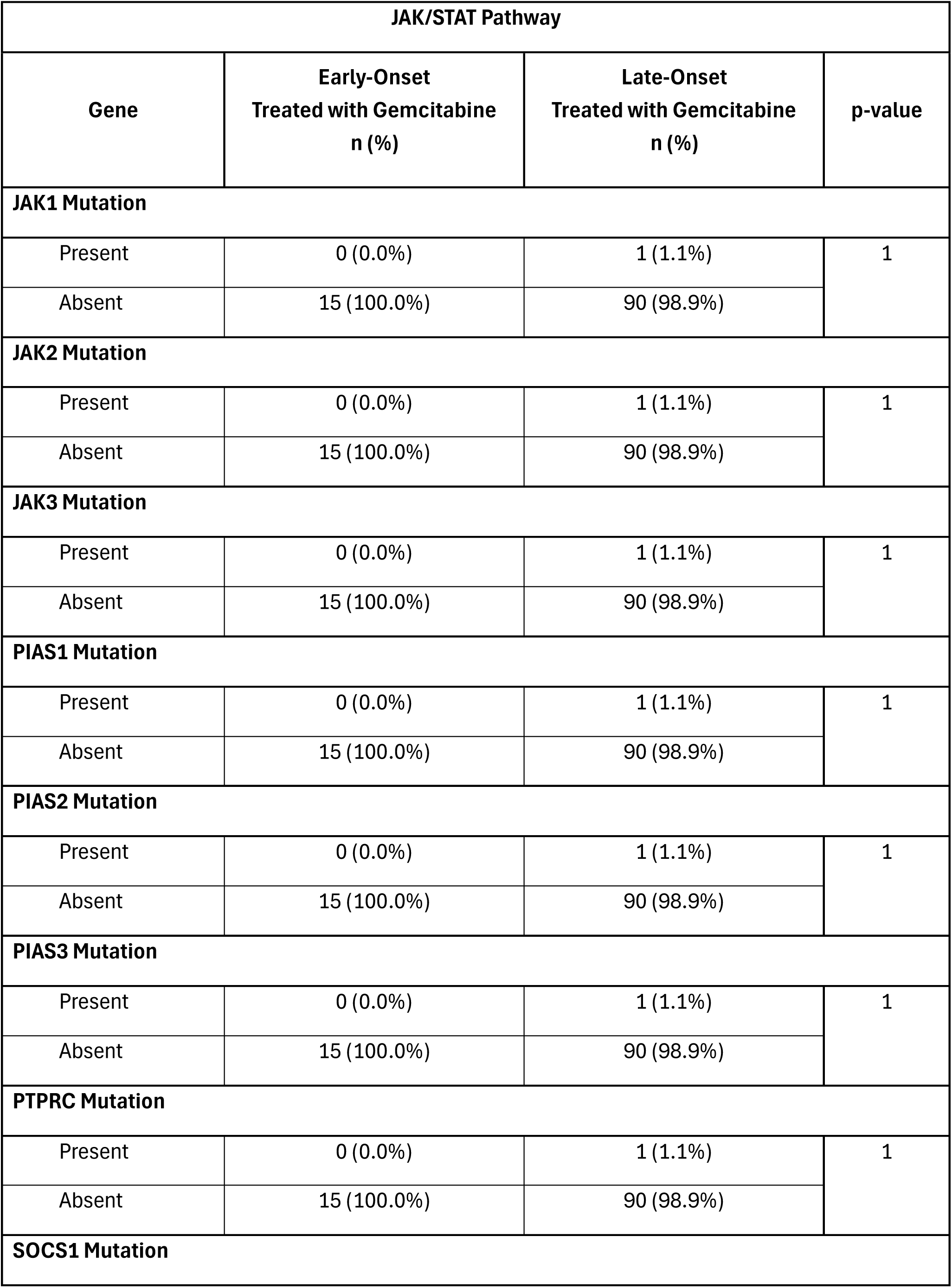
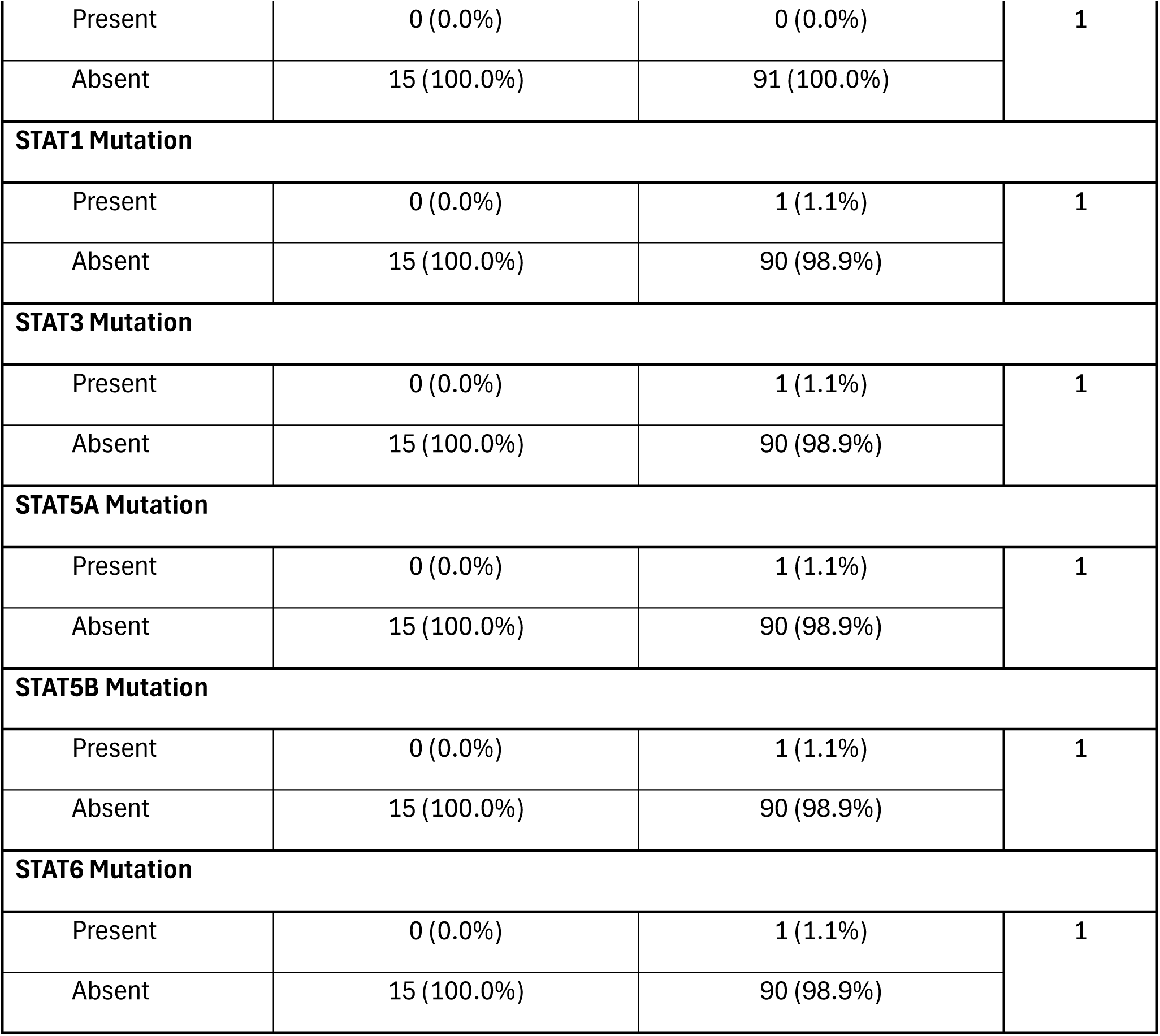
Comparison of Early-Onset PDAC Patients Versus Late-Onset PDAC Patients Treated with Gemcitabine.

| Gene | Early-Onset<br>Treated with Gemcitabine<br>n (%) | Late-Onset<br>Treated with Gemcitabine<br>n (%) | p-value |
| --- | --- | --- | --- |
| JAK1 Mutation |  |  |  |
| Present | 0 (0.0%) | 1 (1.1%) | 1 |
| Absent | 15 (100.0%) | 90 (98.9%) |  |
| JAK2 Mutation |  |  |  |
| Present | 0 (0.0%) | 1 (1.1%) | 1 |
| Absent | 15 (100.0%) | 90 (98.9%) |  |
| JAK3 Mutation |  |  |  |
| Present | 0 (0.0%) | 1 (1.1%) | 1 |
| Absent | 15 (100.0%) | 90 (98.9%) |  |
| PIAS1 Mutation |  |  |  |
| Present | 0 (0.0%) | 1 (1.1%) | 1 |
| Absent | 15 (100.0%) | 90 (98.9%) |  |
| PIAS2 Mutation |  |  |  |
| Present | 0 (0.0%) | 1 (1.1%) | 1 |
| Absent | 15 (100.0%) | 90 (98.9%) |  |
| PIAS3 Mutation |  |  |  |
| Present | 0 (0.0%) | 1 (1.1%) | 1 |
| Absent | 15 (100.0%) | 90 (98.9%) |  |
| PTPRC Mutation |  |  |  |
| Present | 0 (0.0%) | 1 (1.1%) | 1 |
| Absent | 15 (100.0%) | 90 (98.9%) |  |
| SOCS1 Mutation |  |  |  |
| Present | 0 (0.0%) | 0 (0.0%) | 1 |
| Absent | 15 (100.0%) | 91 (100.0%) |  |
| STAT1 Mutation |  |  |  |
| Present | 0 (0.0%) | 1 (1.1%) | 1 |
| Absent | 15 (100.0%) | 90 (98.9%) |  |
| STAT3 Mutation |  |  |  |
| Present | 0 (0.0%) | 1 (1.1%) | 1 |
| Absent | 15 (100.0%) | 90 (98.9%) |  |
| STAT5A Mutation |  |  |  |
| Present | 0 (0.0%) | 1 (1.1%) | 1 |
| Absent | 15 (100.0%) | 90 (98.9%) |  |
| STAT5B Mutation |  |  |  |
| Present | 0 (0.0%) | 1 (1.1%) | 1 |
| Absent | 15 (100.0%) | 90 (98.9%) |  |
| STAT6 Mutation |  |  |  |
| Present | 0 (0.0%) | 1 (1.1%) | 1 |
| Absent | 15 (100.0%) | 90 (98.9%) |  |

**Table S8.** Comparison of Early-Onset PDAC Patients Versus Late-Onset PDAC Patients Not Treated with Gemcitabine.

| **JAK/STAT Pathway** |  |  |  |
| --- | --- | --- | --- |
| **Gene** | **Early-Onset**<br>**Not Treated with**<br>**Gemcitabine**<br>**n (%)** | **Late-Onset**<br>**Not Treated with**<br>**Gemcitabine**<br>**n (%)** | **p-value** |
| **JAK1 Mutation** |  |  |  |
| Present | 0 (0.0%) | 1 (1.1%) | 1 |
| Absent | 5 (100.0%) | 90 (98.9%) |  |
| JAK2 Mutation |  |  |  |
| Present | 0 (0.0%) | 1 (1.1%) | 1 |
| Absent | 5 (100.0%) | 90 (98.9%) |  |
| JAK3 Mutation |  |  |  |
| Present | 0 (0.0%) | 1 (1.1%) | 1 |
| Absent | 5 (100.0%) | 90 (98.9%) |  |
| PIAS1 Mutation |  |  |  |
| Present | 0 (0.0%) | 1 (1.1%) | 1 |
| Absent | 5 (100.0%) | 90 (98.9%) |  |
| PIAS2 Mutation |  |  |  |
| Present | 0 (0.0%) | 1 (1.1%) | 1 |
| Absent | 5 (100.0%) | 90 (98.9%) |  |
| PIAS3 Mutation |  |  |  |
| Present | 0 (0.0%) | 1 (1.1%) | 1 |
| Absent | 5 (100.0%) | 90 (98.9%) |  |
| PTPRC Mutation |  |  |  |
| Present | 0 (0.0%) | 1 (1.1%) | 1 |
| Absent | 5 (100.0%) | 90 (98.9%) |  |
| SOCS1 Mutation |  |  |  |
| Present | 0 (0.0%) | 0 (0.0%) | 1 |
| Absent | 5 (100.0%) | 91 (100.0%) |  |
| STAT1 Mutation |  |  |  |
| Present | 0 (0.0%) | 1 (1.1%) | 1 |
| Absent | 5 (100.0%) | 90 (98.9%) |  |
| STAT3 Mutation |  |  |  |
| Present | 0 (0.0%) | 1 (1.1%) | 1 |
| Absent | 5 (100.0%) | 90 (98.9%) |  |
| STAT5A Mutation |  |  |  |
| Present | 0 (0.0%) | 1 (1.1%) | 1 |
| Absent | 5 (100.0%) | 90 (98.9%) |  |
| STAT5B Mutation |  |  |  |
| Present | 0 (0.0%) | 1 (1.1%) | 1 |
| Absent | 5 (100.0%) | 90 (98.9%) |  |
| STAT6 Mutation |  |  |  |
| Present | 0 (0.0%) | 1 (1.1%) | 1 |
| Absent | 5 (100.0%) | 90 (98.9%) |  |

**Figure S2.**
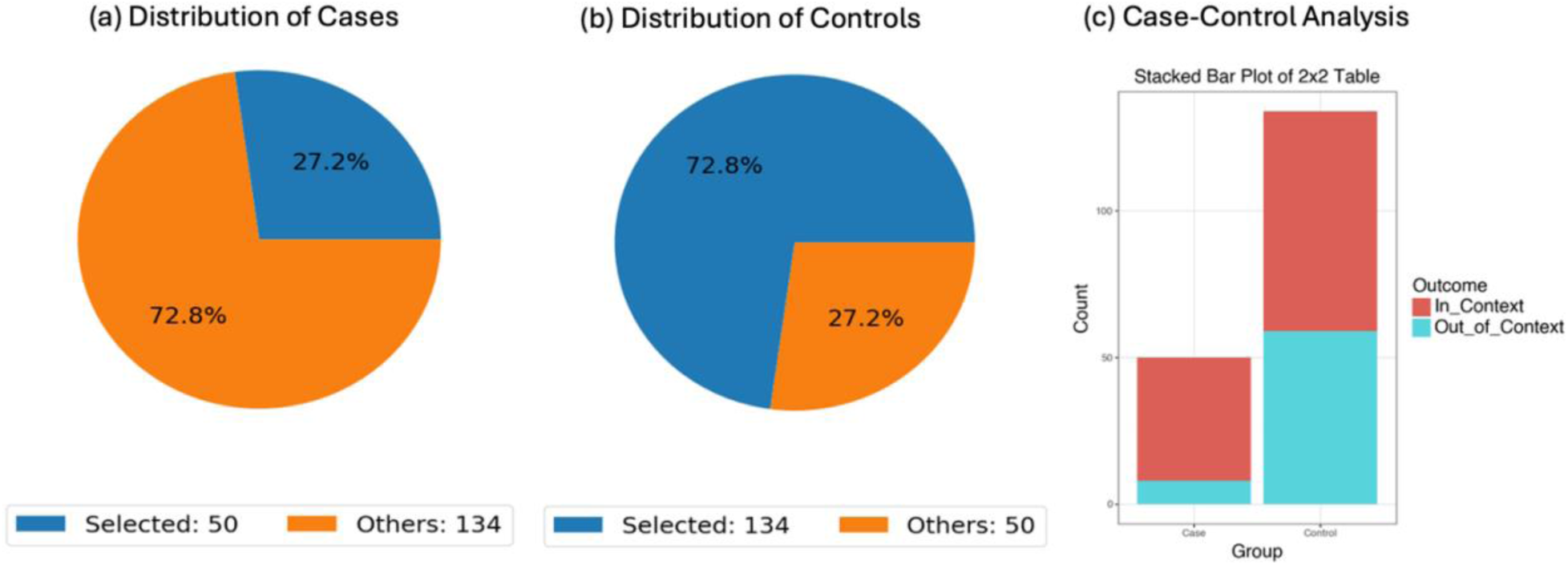
Conversational AI–assisted enrichment analysis of KRAS mutations according to TGFβ pathway status in PDAC. This figure illustrates a case-control analysis performed using the AI-HOPE-TGFβ conversational artificial intelligence framework to determine whether KRAS mutations are enriched among pancreatic ductal adenocarcinoma (PDAC) tumors harboring TGFβ pathway alterations. Using natural language–driven cohort selection, the framework identified (a) a case cohort consisting of tumors with TGFβ pathway alterations (n = 50; 27.2% of the study population) and (b) a control cohort comprising tumors without detectable TGFβ pathway alterations (n = 134; 72.8%). Pie charts depict the relative distribution of selected and non-selected samples within each cohort. (c) A stacked bar plot summarizes the frequency of KRAS-mutant and KRAS–non-mutant tumors across case and control groups. Statistical evaluation using Fisher’s exact test demonstrated a significant association between KRAS mutation status and TGFβ pathway alterations, yielding an odds ratio of 4.13 (95% CI: 1.80–9.47; p = 0.001). These findings suggest that tumors with TGFβ pathway dysregulation are substantially more likely to harbor KRAS mutations, supporting the biological interplay between KRAS-driven oncogenic signaling and TGFβ-mediated tumor progression in PDAC. Furthermore, this analysis highlights the ability of conversational AI agents to rapidly perform interpretable enrichment analyses and uncover clinically relevant molecular associations within complex cancer genomic datasets.

**Figure S3.**
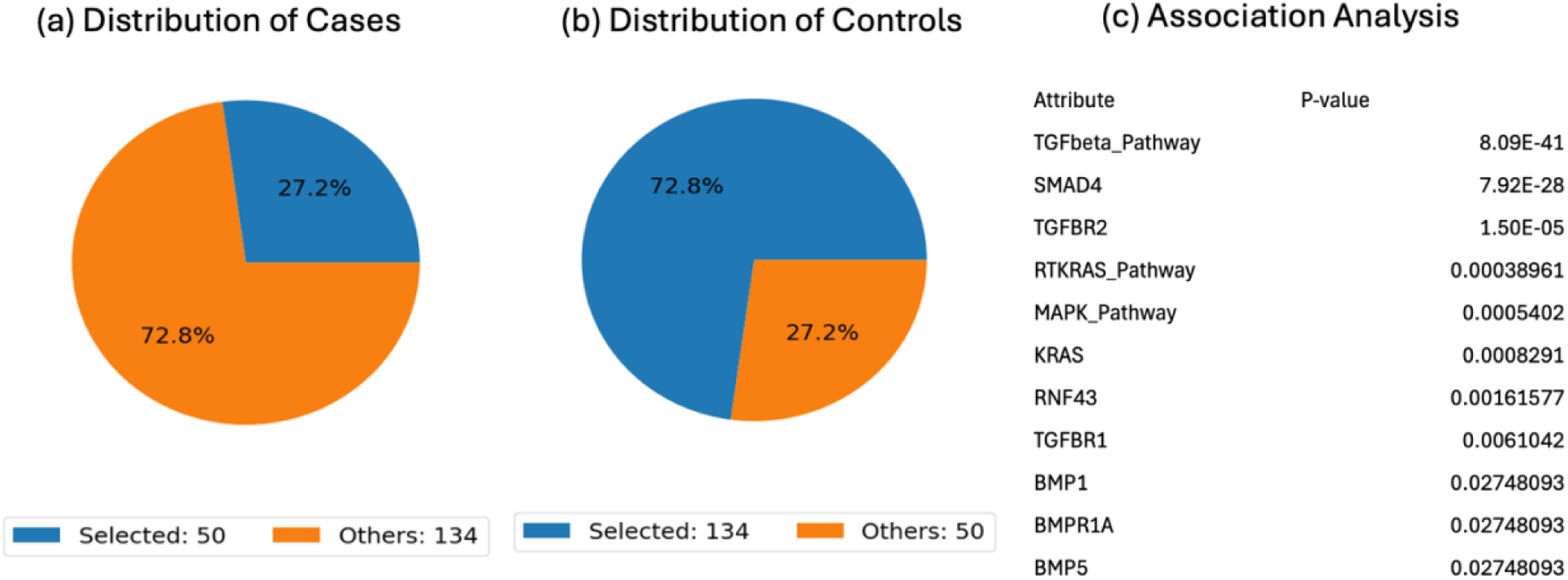
Conversational AI–enabled discovery of clinicogenomic associations linked to TGFβ pathway alterations in PDAC. This figure summarizes a comprehensive association analysis performed using the AI-HOPE-TGFβ conversational artificial intelligence framework to identify molecular and clinical characteristics associated with TGFβ pathway dysregulation in pancreatic ductal adenocarcinoma (PDAC). The case cohort consisted of tumors harboring TGFβ pathway alterations (n = 50; 27.2%), while the control cohort included tumors lacking detectable TGFβ pathway alterations (n = 134; 72.8%). Panels (a) and (b) display the proportional distribution of selected and non-selected samples within the case and control populations, respectively. Panel (c) presents the results of a large-scale association analysis comparing TGFβ pathway status with clinicogenomic variables across the cohort. As expected, the strongest associations were observed with canonical TGFβ signaling components, including SMAD4, TGFBR2, and TGFBR1, highlighting the central role of these genes in defining pathway dysregulation. Significant relationships were also identified with broader oncogenic signaling networks, including alterations in the RTK-RAS and MAPK pathways, as well as KRAS, the dominant driver mutation in PDAC. Additional associations involving RNF43 and members of the bone morphogenetic protein (BMP) signaling family (BMP1, BMPR1A, and BMP5) further suggest coordinated perturbation of interconnected developmental and growth factor signaling pathways. These findings indicate that TGFβ pathway-altered tumors represent a distinct molecular subset of PDAC characterized by concurrent abnormalities in KRAS-driven oncogenic signaling and multiple components of the TGFβ/BMP regulatory network. This analysis demonstrates the ability of conversational artificial intelligence agents to rapidly identify biologically meaningful associations and uncover complex pathway-level interactions within multidimensional cancer genomic datasets, thereby supporting precision medicine–oriented hypothesis generation and biomarker discovery.

**Figure S4.**
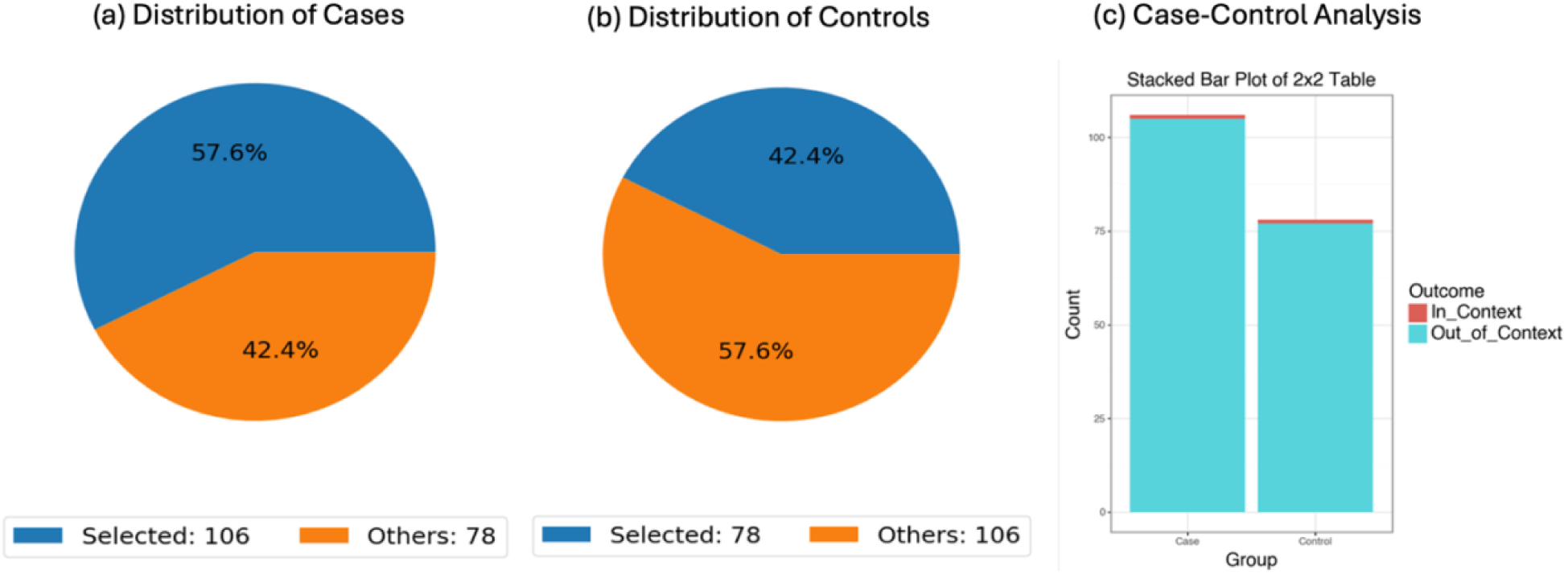
Conversational AI–driven comparison of STAT1 mutation prevalence between gemcitabine-treated and non-gemcitabine-treated PDAC patients. This figure presents an odds ratio analysis performed using the AI-HOPE platform to determine whether STAT1 mutation frequency differs according to gemcitabine treatment exposure in pancreatic ductal adenocarcinoma (PDAC). The case cohort consisted of patients who received gemcitabine-based therapy (n = 106), while the control cohort comprised patients who did not receive gemcitabine (n = 78). Panels (a) and (b) illustrate the relative representation of the treatment-defined cohorts within the overall study population, highlighting the larger proportion of gemcitabine-treated patients. Panel (c) summarizes the distribution of STAT1-mutant and STAT1–wild-type tumors between the two treatment groups. STAT1 mutations were identified in 1 of 106 (0.94%) gemcitabine-treated patients and 1 of 78 (1.28%) non-gemcitabine-treated patients. Statistical analysis demonstrated no significant difference in STAT1 mutation frequency between treatment groups (Fisher’s exact test p = 1.0). The estimated odds ratio was 0.73 (95% CI: 0.045–11.91), indicating no evidence of enrichment or depletion of STAT1 mutations among patients receiving gemcitabine. These findings suggest that genomic alterations affecting STAT1 are exceedingly rare in PDAC and do not appear to be associated with gemcitabine treatment status. Furthermore, this analysis illustrates the ability of conversational artificial intelligence agents to rapidly construct clinically relevant cohorts, perform automated statistical comparisons, and identify treatment-associated molecular patterns within precision oncology datasets.

**Figure S5.**
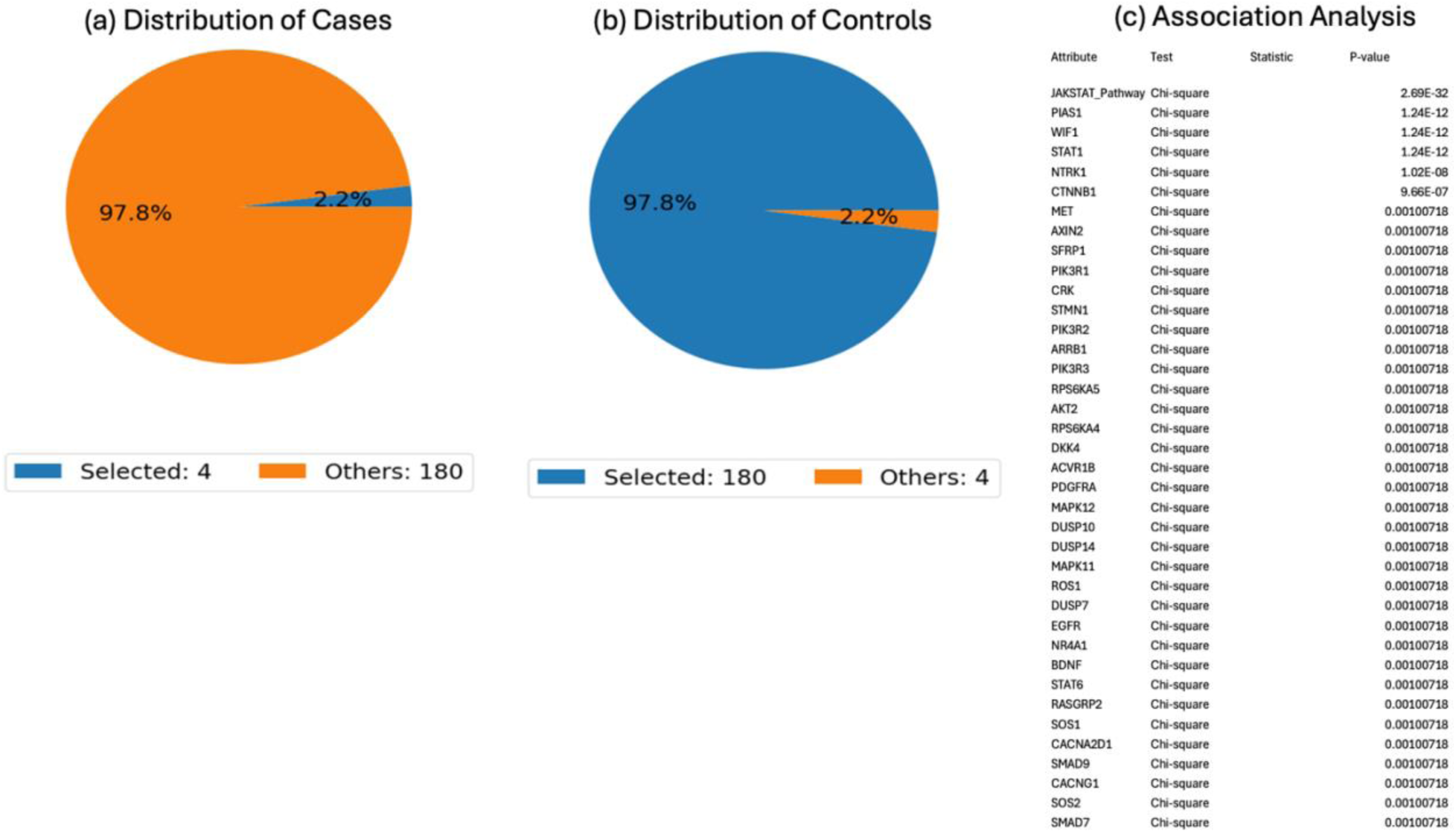
AI-guided identification of genomic correlates of JAK/STAT pathway dysregulation in pancreatic ductal adenocarcinoma. This figure presents a conversational artificial intelligence–driven association analysis designed to characterize the molecular landscape associated with JAK/STAT pathway alterations in PDAC. Using the AI-HOPE-JAK/STAT framework, tumors harboring JAK/STAT pathway alterations were designated as the case cohort (n = 4; 2.2% of the cohort), while tumors lacking detectable pathway alterations served as controls (n = 180; 97.8%). Panels (a) and (b) illustrate the distribution of pathway-altered and pathway-unaltered tumors within the study population, demonstrating the low prevalence of JAK/STAT pathway alterations relative to the overall cohort. Panel (c) summarizes statistically significant associations identified between JAK/STAT pathway status and clinicogenomic features. The strongest relationships were observed with core JAK/STAT signaling components, including PIAS1, STAT1, and STAT6, validating the biological coherence of the pathway-based classification. Significant associations were also identified with multiple regulators of receptor tyrosine kinase signaling and intracellular signal transduction, including NTRK1, MET, EGFR, PDGFRA, and several members of the PI3K/AKT signaling network (PIK3R1, PIK3R2, PIK3R3, and AKT2). Additional enrichment of genes involved in WNT signaling and transcriptional regulation, such as CTNNB1, WIF1, AXIN2, and SFRP1, suggests extensive cross-talk between JAK/STAT and developmental signaling pathways. Notably, significant associations with SMAD7, SMAD9, and ACVR1B indicate potential interactions between JAK/STAT and TGFβ-related signaling networks, highlighting the interconnected nature of oncogenic pathways in PDAC. Collectively, these findings suggest that tumors with JAK/STAT pathway alterations represent a rare but biologically distinctive subgroup characterized by coordinated perturbations across growth factor, PI3K/AKT, MAPK, WNT, and TGFβ signaling pathways. This analysis further demonstrates the utility of conversational artificial intelligence agents for rapidly uncovering multidimensional molecular relationships and generating mechanistically relevant hypotheses within precision oncology datasets.

**Figure S6.**
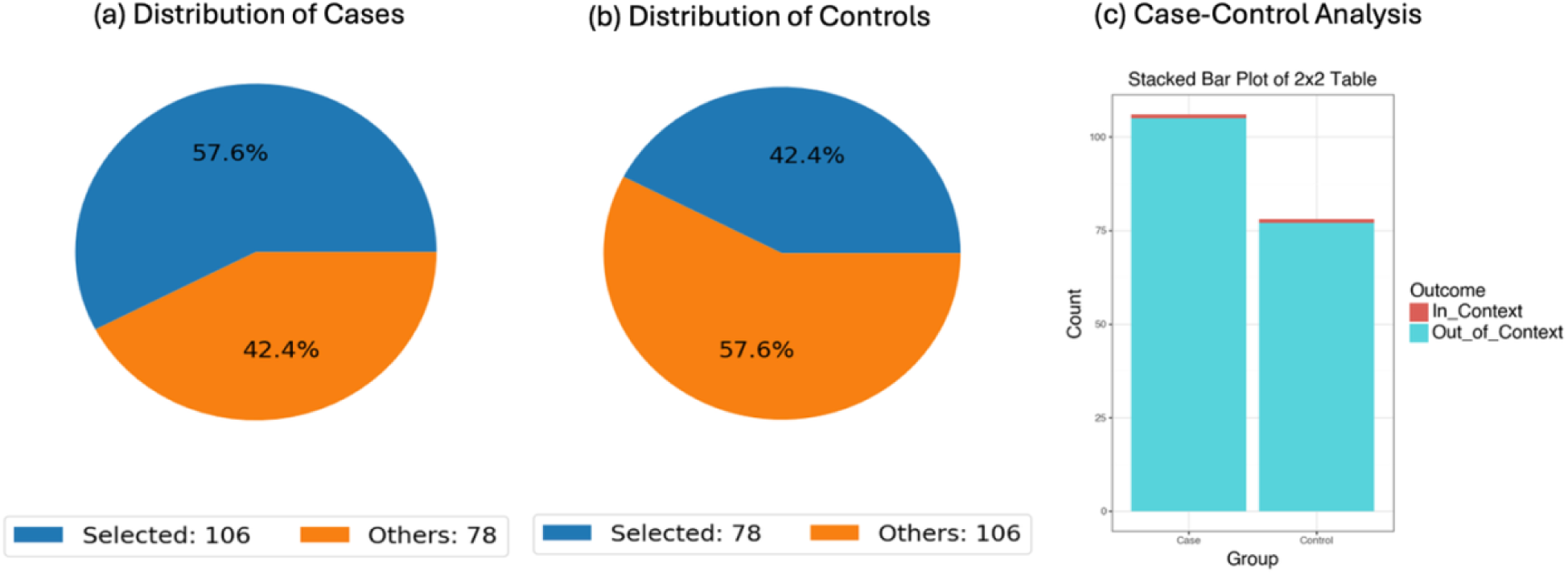
Conversational AI–assisted assessment of STAT1 mutation frequency according to gemcitabine treatment status in PDAC. This figure presents a case-control analysis performed using the AI-HOPE-JAK/STAT framework to determine whether STAT1 mutations are differentially distributed between pancreatic ductal adenocarcinoma (PDAC) patients who received gemcitabine-based therapy and those who did not. The case cohort consisted of gemcitabine-treated patients (n = 106; 57.6% of the cohort), while the control cohort included patients without documented gemcitabine exposure (n = 78; 42.4%). Panels (a) and (b) display the relative proportions of selected and non-selected samples within each treatment group. Panel (c) summarizes the distribution of STAT1-mutant and STAT1–wild-type tumors across the two cohorts. STAT1 mutations were uncommon in both groups, occurring in 0.94% of gemcitabine-treated patients and 1.28% of non-treated patients. Statistical comparison using Fisher’s exact test revealed no significant association between gemcitabine exposure and STAT1 mutation status (p = 1.0). The estimated odds ratio was 0.73 (95% CI: 0.045–11.91), indicating no evidence of enrichment or depletion of STAT1 mutations among patients receiving gemcitabine. These findings suggest that STAT1 genomic alterations are rare events in PDAC and are not preferentially associated with gemcitabine treatment status within this cohort. More broadly, this analysis demonstrates the ability of conversational artificial intelligence agents to rapidly perform treatment-stratified genomic comparisons and identify clinically relevant molecular patterns within precision oncology datasets.

